# Microbiome-based risk prediction in incident heart failure: a community challenge

**DOI:** 10.1101/2023.10.12.23296829

**Authors:** Pande Putu Erawijantari, Ece Kartal, José Liñares-Blanco, Teemu D. Laajala, Lily Elizabeth Feldman, The FINRISK Microbiome DREAM Challenge and ML4Microbiome Communities, Pedro Carmona-Saez, Rajesh Shigdel, Marcus Joakim Claesson, Randi Jacobsen Bertelsen, David Gomez-Cabrero, Samuel Minot, Jacob Albrecht, Verena Chung, Michael Inouye, Pekka Jousilahti, Jobst-Hendrik Schultz, Hans-Christoph Friederich, Rob Knight, Veikko Salomaa, Teemu Niiranen, Aki S. Havulinna, Julio Saez-Rodriguez, Rebecca T. Levinson, Leo Lahti

## Abstract

Heart failure (HF) is a major public health problem. Early identification of at-risk individuals could allow for interventions that reduce morbidity or mortality. The community-based FINRISK Microbiome DREAM challenge (synapse.org/finrisk) evaluated the use of machine learning approaches on shotgun metagenomics data obtained from fecal samples to predict incident HF risk over 15 years in a population cohort of 7231 Finnish adults (FINRISK 2002, n=559 incident HF cases). Challenge participants used synthetic data for model training and testing. Final models submitted by seven teams were evaluated in the real data. The two highest-scoring models were both based on Cox regression but used different feature selection approaches. We aggregated their predictions to create an ensemble model. Additionally, we refined the models after the DREAM challenge by eliminating phylum information. Models were also evaluated at intermediate timepoints and they predicted 10-year incident HF more accurately than models for 5- or 15-year incidence. We found that bacterial species, especially those linked to inflammation, are predictive of incident HF. This highlights the role of the gut microbiome as a potential driver of inflammation in HF pathophysiology. Our results provide insights into potential modeling strategies of microbiome data in prospective cohort studies. Overall, this study provides evidence that incorporating microbiome information into incident risk models can provide important biological insights into the pathogenesis of HF.

## 1. Introduction

Cardiovascular diseases (CVD) are the leading cause of death worldwide ^1^. Despite significant advancements in medical therapies, heart failure (HF), a heterogeneous condition characterized by the heart’s inability to pump a sufficient supply of blood, remains a common manifestation of CVD. According to recent estimates, more than 64 million people worldwide have HF and the average 5-year mortality rate worldwide is estimated to be over 55% ^2^. Many HF patients are diagnosed during acute care despite the presence of preceding symptoms ^3^. The early identification of individuals at risk prior to HF onset could allow for early intervention and the subsequent reduction of adverse events including hospitalization, morbidity, and mortality.

Analysis of multiple markers across various omics platforms has provided new opportunities to investigate the underlying factors and physiological processes involved in HF, particularly in the context of exploring potential biomarkers ^4^. In particular, the human gut microbiome, which comprises trillions of microorganisms residing in the gastrointestinal tract, is increasingly recognized as an important modulator of various aspects of human health and disease ^5–10^. The gut microbiome has been suggested as crucial in the pathophysiology of HF ^11–14^, where it has been shown to impact several mechanisms involved in development and progression, including inflammation, oxidative stress, and endothelial dysfunction ^15,16^. While associations between gut microbiome composition and HF have been reported in cross-sectional studies ^14,17,18^, prospective population-level studies on the gut microbiome and incident HF have been lacking. An improved understanding of these prospective associations can support early identification of high-risk individuals.

Community challenges have proven to be a successful strategy for assessing the solvability of bioinformatic problems, identifying best practices, and optimizing predictive features and machine learning models ^19,20^. The recently held Preterm Birth Microbiome Prediction DREAM Challenge ^20^ focused on utilizing vaginal microbiome data from nine studies to predict both preterm and early preterm births. The prominent predictive factors encompassed alpha diversity, community state types, and composition. To further explore the potential of microbiome data in predictive health research, we implemented the Heart Failure Prediction: Microbiome-FINRISK DREAM Challenge to use a community challenge approach to investigate whether the microbiome can predict HF incidence in a prospective population survey of Finnish adults (FINRISK 2002) ^21^.

This challenge provides an avenue for evaluating different modeling approaches of microbiome data in incident risk predictions. Overall, the best-performing models identified during the challenge highlight strategies for incorporating microbiome data in HF risk prediction and provide insights into the possible connections between the gut microbiome and HF. Notably, the co-abundance network approach outperformed other microbiome feature engineering techniques. Taken together, these findings suggest that microbiome data may offer added value to complement conventional risk factors and aid in identifying individuals with a high risk of developing HF.

## 2. Results

### 2.1 Baseline characteristics and heart failure incidence

Of the 7231 individuals in FINRISK 2002 (Figure 1A), 162 (2.2%) had HF at baseline, and another 559 (7.73%) experienced HF during the ∼15-year follow-up period. The mean age of the population was 49.5±13.0 years and 44.9% were men (Table 1). The dataset was randomly divided into three sets: training (n=3615, 50%, n HF=275), testing (n=1807, 25%, n HF=131), and scoring set (n=1809, 25%, n HF=153) (Figure 1C). We confirmed that these groups had a similar microbial taxonomic composition (Figure 1B).

**Figure 1.**
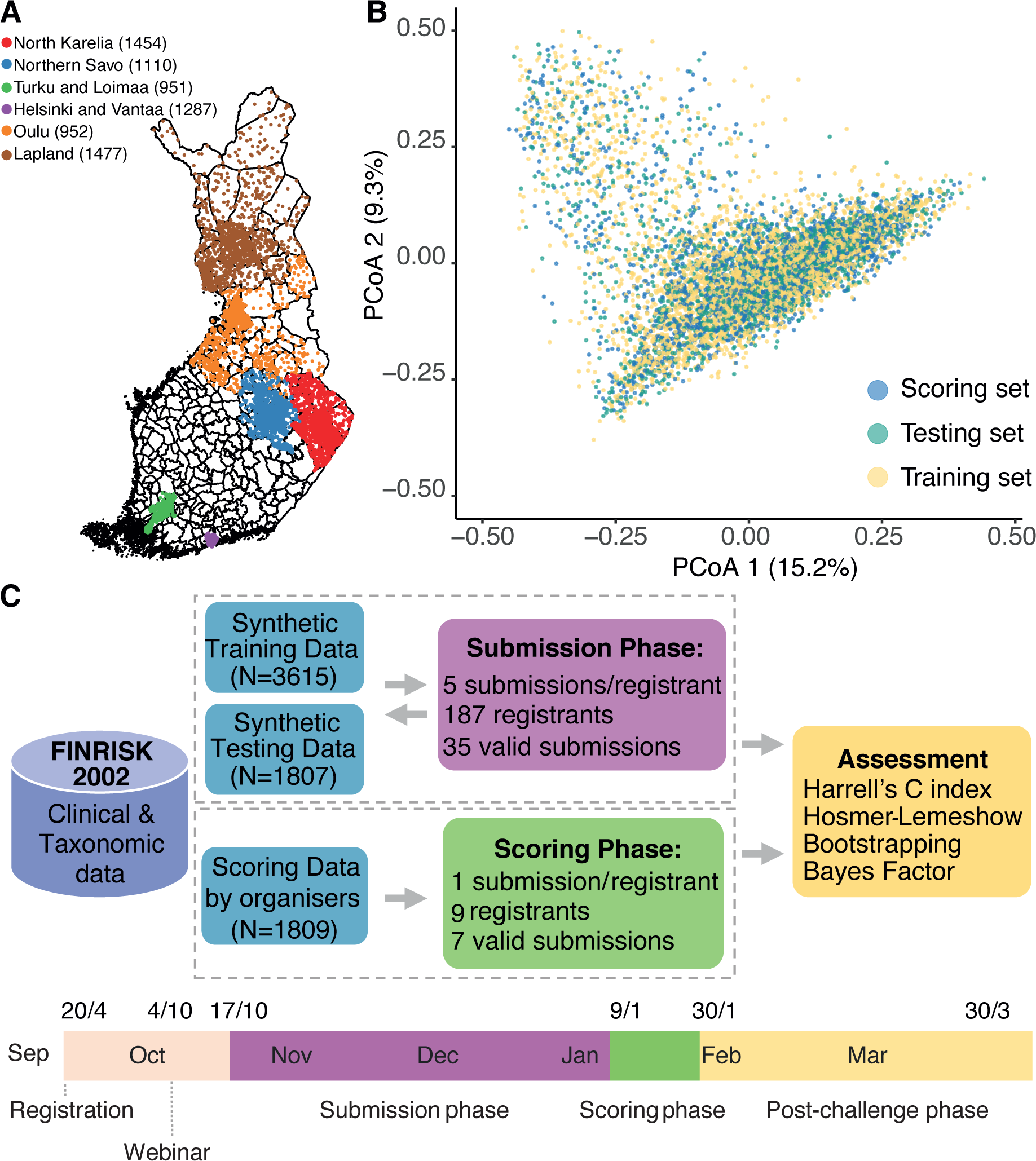
Overview of the DREAM Challenge and FINRISK data. **A.** Geographical distribution across Finland for the individuals within the national FINRISK 2002 cohort. **B.** Principal Coordinate Analysis (PCoA) using Bray-Curtis dissimilarity metrics between randomly selected subsets of the data (training, testing, scoring sets). **C.** The setup and timeline of the DREAM Challenge including submission and scoring phases.

**Table 1.** Overview of the National FINRISK 2002 Cohort.

| Variable | Total | Incident Heart Failure |  | P-value* |
| --- | --- | --- | --- | --- |
|  |  | Yes | No |  |
| **Number** | 7231 | 559 | 6556 |  |
| **Men** | 3248 (44.9%) | 293 (52.4%) | 2909 (44.4%) | 0.00028 |
| **Baseline age, years** | 49.5 (13) | 61.9 (9) | 48.4 (12.7) | 4.2e-125 |
| **Body Mass Index, kg/m<sup>2</sup>** | 27.01 (4.69) | 29.69 (5.05) | 26.78 (4.58) | 2.7e-43 |
| **Systolic Blood Pressure, mmHg** | 135.81 (20.33) | 149.22 (21.93) | 134.69 (19.71) | 9.9e-54 |
| **Non HDL Cholesterol, mmol/L** | 4.09 (1.09) | 4.29 (1.07) | 4.07 (1.08) | 3.8e-06 |
| **Current smokers** | 1687 (23.3%) | 113 (20.2%) | 1545 (23.6%) | 0.085 |
| **Taking Blood Pressure Treatment** | 1129 (15.6%) | 213 (38.1%) | 900 (13.7%) | 6.1e-42 |
| **Diabetes at baseline** | 407 (5.6%) | 85 (15.2%) | 322 (4.9%) | 6.2e-18 |
| **Coronary Heart Diseases at baseline** | 205 (2.8%) | 48 (8.6%) | 157 (2.4%) | 2.3e-12 |
| **Heart Failure at baseline** | 162 (2.2%) | NA | NA | NA |

Data are presented as n (%) (n of participants in indicated category and percentage of total) or mean (SD).
\*Mann-Whitney U test was used for numeric data; Fisher exact test was used for categorical data.

### 2.2. DREAM challenge model performance

To identify optimal modeling strategies for microbiome-based risk prediction in prospective cohort studies using machine learning (ML) approaches, we designed and initiated an open DREAM community challenge (Figure 1C, https://www.synapse.org/finrisk). The challenge participants developed their prediction models using the synthetic training dataset (Figure 1C, Supp. Figure 1A-B, Supp. Table 1). During the submission phase, participants could submit up to five models for hyperparameter tuning, which were conducted based on feedback provided to participants as challenge organizers evaluated their models on real testing datasets. The final models were then submitted and the challenge organizers evaluated them in the real scoring data set during the scoring phase. There were 187 registered participants and ten teams submitted models in the submission phase (Supp. Table 2) and seven teams submitted final models for evaluation (Figure 1, Supp. Table 3). The performance of the final models was evaluated during the scoring phase in a real FINRISK scoring dataset that was not accessible to participants. Harrell’s C-index was comparable between the four highest-scoring teams in the scoring phase (Supp. Table 3). Out of these, two models achieved calibration comparable to our baseline models (Figure 2, Supp. Figure 2).

**Figure 2.**
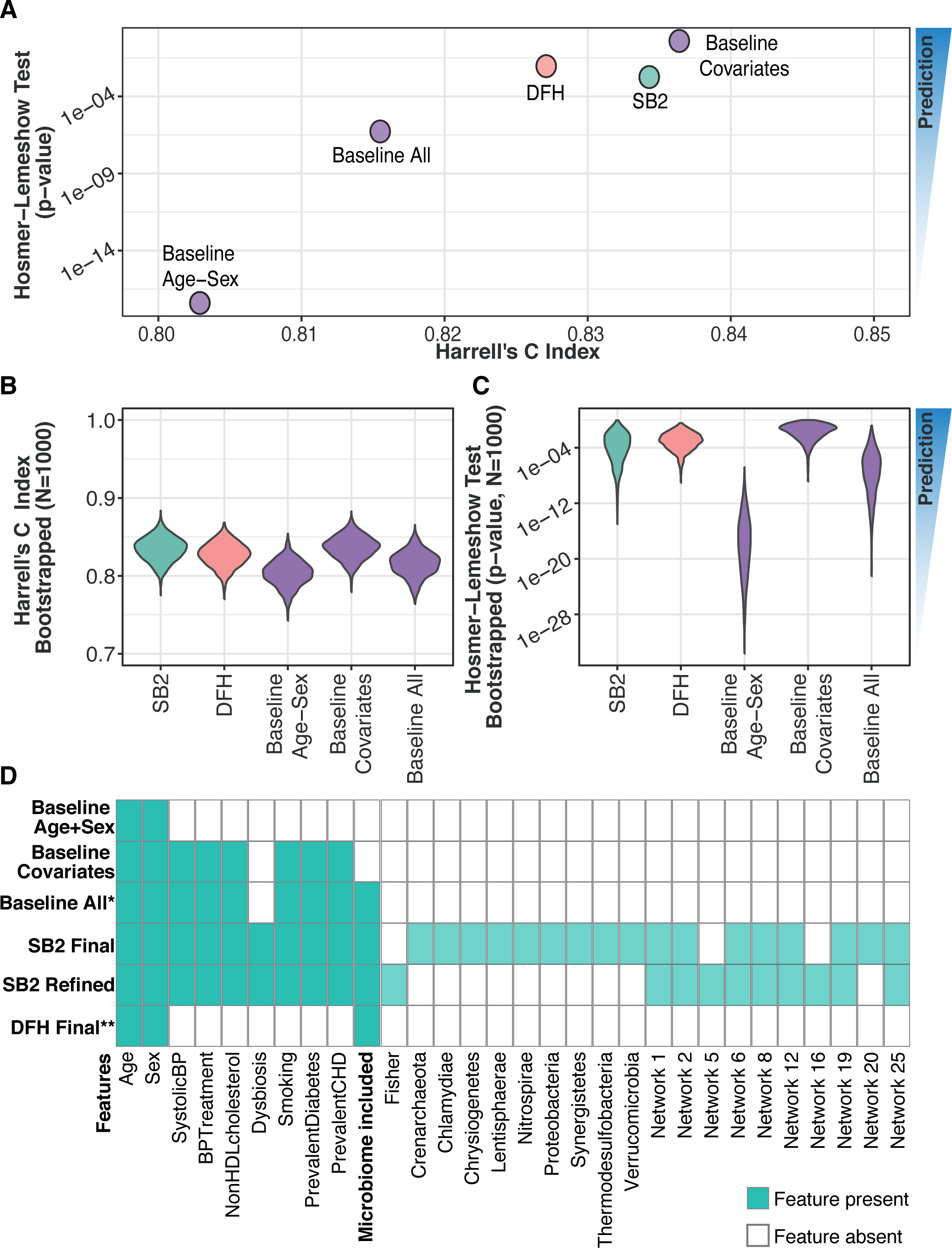
Harrell’s C and Hosmer-Lemeshow test **A.** Harrell’s C-index and Hosmer-Lemeshow p-value were obtained for the investigated models, including the three baseline models provided by the organizers in the scoring phase. **B-C.** Harrell’s C-index and Hosmer-Lemeshow empirical p-value on 1000 bootstrapped iterations for all the models. We used blue for SB2, orange for DFH and purple for the baseline models. **D.** Selected features in the baseline and top models. *Taxonomic features in the “Baseline All” model are presented in Supp. Table 6. ** The features and modules selected by DFH model were weight-based from 10 different seeds. Features present in each model are represented by turquoise-filled squares, while absence is indicated by blank squares.

A schematic overview of the two top-performing models is provided in Figure 3. Both best-scoring models used variations of a regularized Cox proportional hazards model. DenverFINRISKHacky (DFH) team’s final model achieved a Harrell’s C-index of 0.8271 and SB2 team’s model performed slightly better, yielding a Harrell’s C-index of 0.8344 (Figure 2A, Supp. Table 3). The most informative variables for both models are presented in Fig 2D and Supp. Table 4.

**Figure 3.**
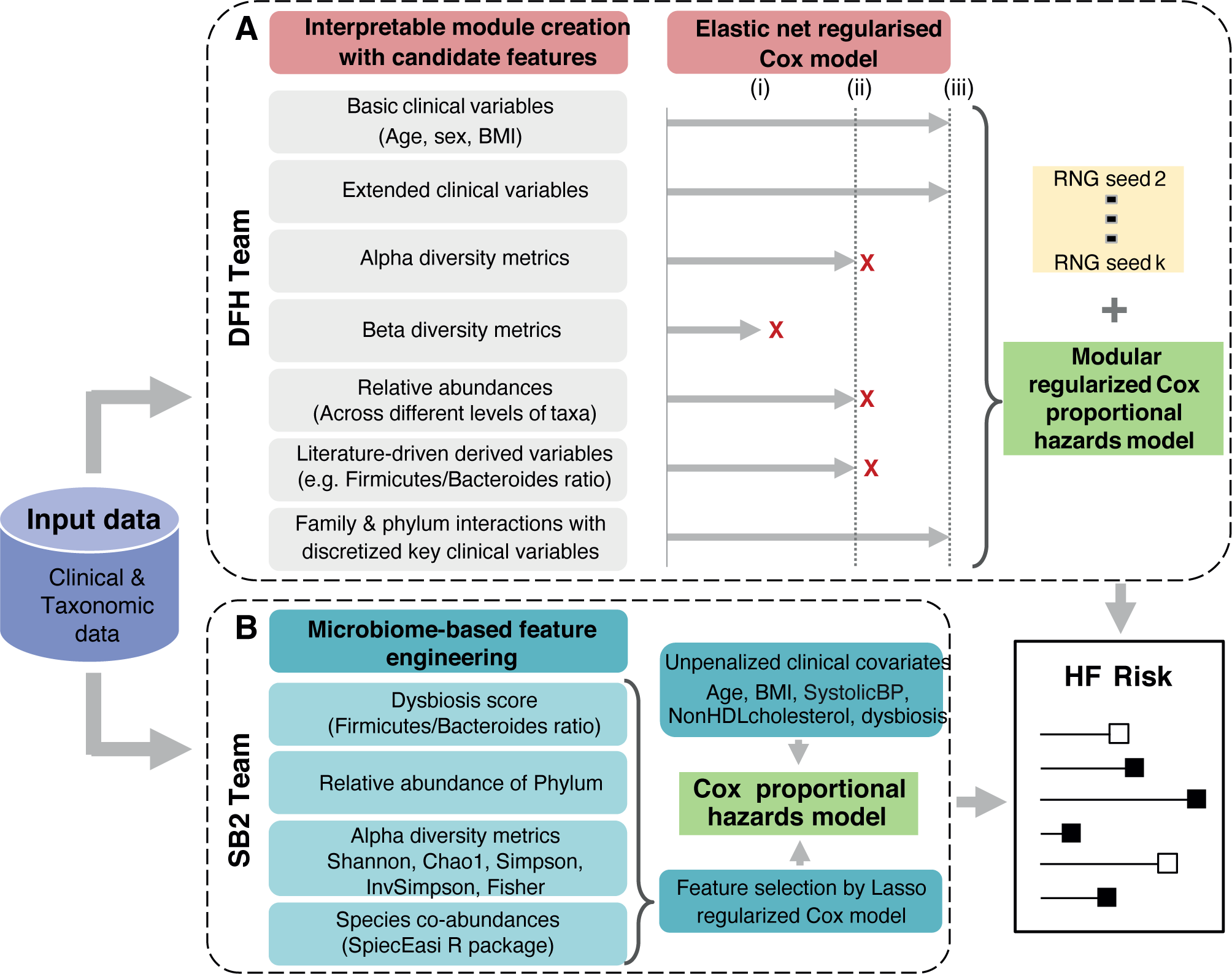
Schematic illustration of modeling workflow of the two top-performing teams. **A.** Team DFH used modular Elastic Net regularized Cox proportional hazards model. After manually curating inter-pretable modules, they identified the optimal features within each module by module-specific cross-validation. The pruned modules were then combined and used to identify the best overall combination of features using cross-validation. The team averaged final risk predictions across multiple seeds. **B.** The SB2 team used LASSO regularization to retain 29 features encompassing age, BMI, systolic blood pressure, non-HDL cholesterol, sex, and dysbiosis as unpenalized features, and blood pressure treatment, prevalent diabetes, smoking and prevalent coronary heart disease were penalized and selected by LASSO to be included in the final Cox proportional hazards model..

The DFH model provided limited ability to interpret the importance of individual features, due to the implementation of variable transformations and non-linear feature combinations. Therefore, we focused our interrogation of the biology represented by the features in the SB2 model (Fig 2D, Supp. Table 4). The SB2 model selected 11 phyla including Crenarchaeota, Bacteroidetes, Verrucomicrobia, Chrysiogenetes, and Proteobacteria and eight co-abundance networks that were identified as relevant for HF risk prediction (Supp. Table 4-5). Notably, the microbiome features that showed the highest association coefficient with HF were Crenarchaeota, Chrysiogenetes, network 1, and 6 (Supp. Table 5, Supp. Figure 3-4). It should be noted that Crenarchaeota and Chrysiogenetes which have been infrequently observed in gut microbiome studies, were identified with relatively low read counts in our samples (Supp. Figure 5). This underscores the importance of further verifications, especially related to potential limitation that arises when rare taxa are either improperly filtered or when computational methods were employed for microbiome feature selection in data-driven models without considering prior knowledge.

Network 6 exhibited a diverse composition, containing ten species from the class Clostridia, five from Erysipelotrichia, and two from Coriobacteriia (Supp. Table 5). Species from Clostridia class (e.g *Clostridium citroniae, Clostridium asparagiforme, Hungatella hathewayi*) have been linked with the production of trimethylamine N-oxide (TMAO), a compound that is likely connected to the intake of red meat, eggs, and fatty dairy products ^22,23^, and has been previously linked with cardiovascular diseases ^24,25^. Network 6 also contains *Clostridium bolteae* and *Clostridium citroniae,* which has been positively associated with inflammatory signals ^17,26,27^ and associated with type 2 diabetes (T2D) ^28,29^. Additionally, network 6 also contained opportunistic pathogens, such as *Hungatella hathewayi*, *Clostridium ramosum*, *Clostridium symbiosum, and Eggerthella lenta* ^28^. Network 1 (Supp. Table 5) contained 10 species from the class *Coriobacteriia*. Among them, *Collinsella* genera abundance has been associated with T2D ^30^, and in overweight and obese pregnant women ^31^.

### 2.3 Model refinement

As phylum level information introduces redundancy with higher resolution taxonomic level (Supp. Table 4), we also ran the model without phylum information (SB2 refined model). This slightly improved the performance and fit of this model (Harrell’s C-index=0.8388, Hosmer-Lemeshow p-value=0.0102; Supp. Table 7). The relevant co-abundance networks remained similar with additional contributions of network 20 (Supp. Table 5), which also contains *R. gnavus* that has been positively associated with inflammation ^32,33^. In addition, we observed that Fisher’s alpha diversity index contributed to the final model (Fig 2D, Supp. Table 4). Restricting the unpenalized information to age (SB2 age fixed model) provided a modest improvement in model accuracy (Harrell’s C-index=0.8392, Hosmer-Lemeshow p-value= 0.0019; Supp. Table 7). The model selected all clinical covariates and additional co-abundance networks (networks 9, 11, 16 and 24) while removing dysbiosis as a feature. Four co-abundance networks (1, 6, 12, and 25) had positive associations with incident HF (Hazard ratio > 1, Supp. Table 4). All models consistently referred to clinical covariates (age, sex, BMI, smoking status, blood pressure, prevalent diabetes and prevalent coronary heart diseases).

Finally, we investigated whether combining predictions from the top-performing approaches could further improve the accuracy and calibrations of the predictions. Toward this goal, we constructed ensemble models (Figure 4), which were generated by taking the mean of prediction scores obtained from the combined individual models. Mean-aggregated ensemble models of the top 2 performers improved both model performance measured by Harrell’s C-index (Figure 5A, Supp. Table 7; 0.8369) and calibration as determined by Hosmer-Lemeshow test p-value (Figure 4B, Supp. Table 7; 0.1551).

**Figure 4.**
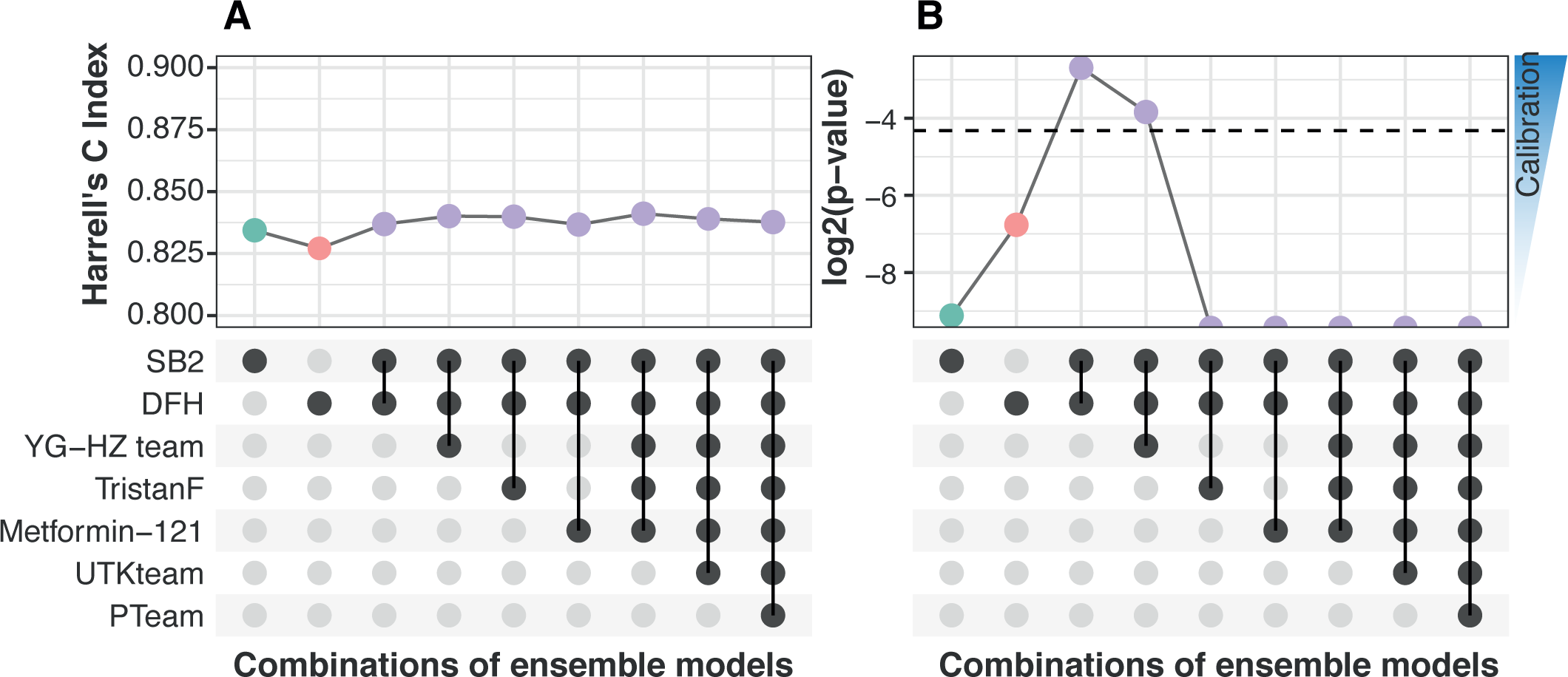
**A.**Harrell’s C-index and **B.** Hosmer-Lemeshow p-value for the ensemble models from mean-aggregations of the final model’s individual risk score. The lower plot illustrates the combination of teams utilized in the calculation of the mean for the aggregated final models. The dashed line corresponds to p-value=0.05 on the y-axis (B), while the x-axis represents different combinations of ensemble models.

**Figure 5.**
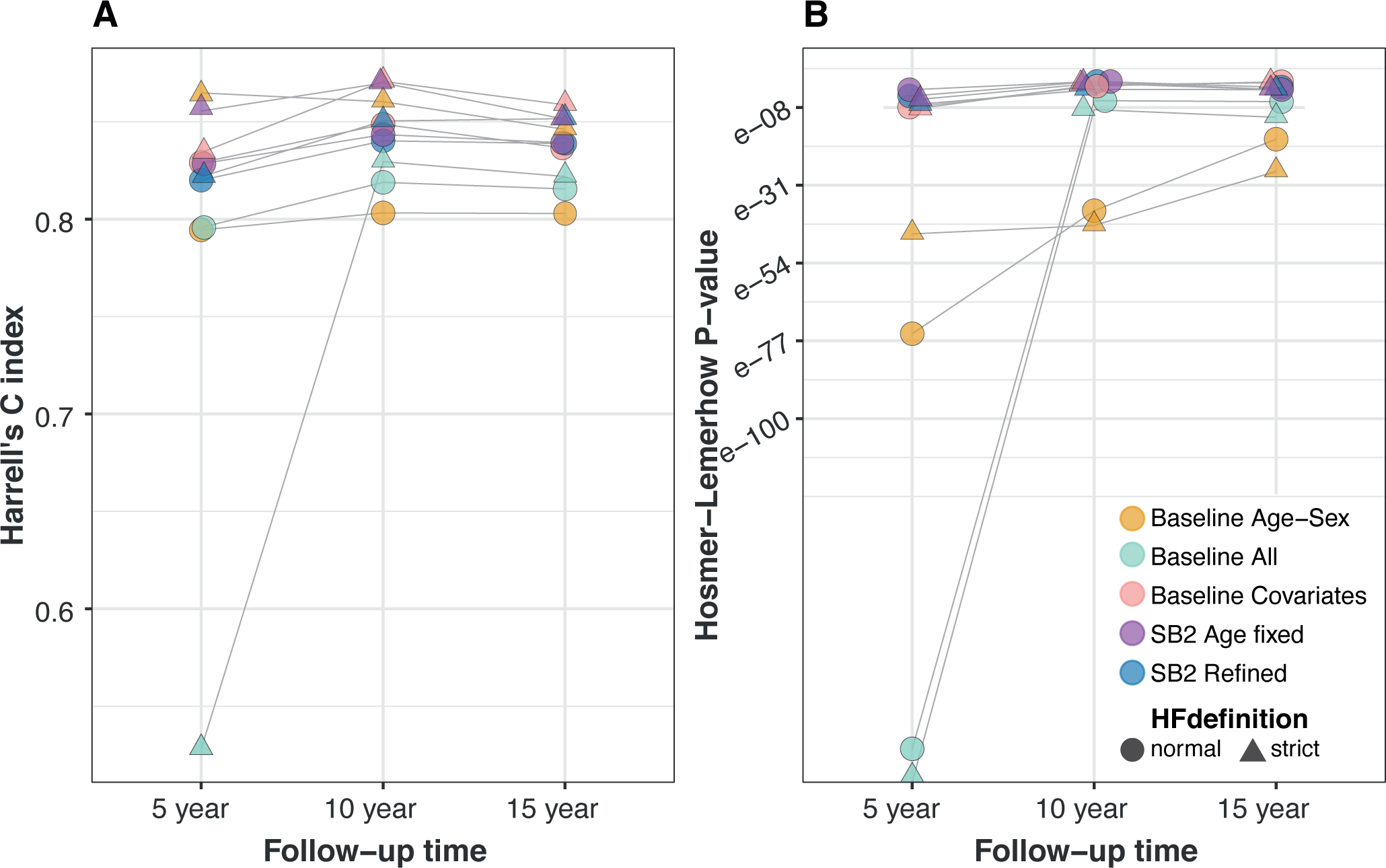
Evaluation of model performance over varying follow-up times. **A.** Harrell’s C-index and **B.** Hosmer-Lemeshow p-values are presented for different models, distinguished by unique colors, across three distinct follow-up periods: 5, 10, and 15 years. Two distinct HF definitions were represented in different shapes.

### 2.4 Enhanced model performance and evaluation results

While the challenge was conducted using a lenient definition of HF and the follow-up endpoint of 15 years, another definition of HF relying on strict criteria, which required additional assessments to confirm the relationship between drug purchases and HF ^34^, was also available in the national FINRISK study. To test whether a population where the incident diagnosis of HF was more certain would further improve predictions with the current models, we tested the models with this stricter definition (N cases=288) ^34^. Both SB2 and DFH models showed improved predictions for the stricter definition of the HF endpoint (C-statistics of 0.8541 and 0.8454 respectively). However, only the SB2 model achieved a well-calibrated performance (Hosmer-Lemeshow p-val=0.1359, Supp. Table 7).

Finally, we evaluated our baseline models and SB2 refined models (SB2 refined and SB2 age fixed models) based on the follow-up times of 5 and 10 years, two intermediate time points that were made available in addition to the original 15-year follow-up period. There were 115 and 299 recorded cases of incident HF within 5 and 10 years of follow-up, respectively. Notably, the 10-year risk prediction models exhibited enhanced performance and improved calibrations, whereas the 5-year follow-up exhibited a decline in performance and calibration when compared to the 15-year follow-up (Figure 5).

## 3. Discussion

This study demonstrates the value of incorporating taxonomic profiling in the prediction of incident HF. We leveraged the National FINRISK 2002 cohort, a unique resource used to identify predictive microbiome signatures for various complex diseases and all-cause mortality^21^. While the model performance did not substantially improve through the inclusion of microbial taxonomic profile on the conventional risk factors, the incorporation of microbiome data did provide important information for biological interpretability. Additionally, incorporation of species-level co-abundance networks improved the performance, as shown by the top-performing model performance compared to the microbiome-based baseline model (Baseline all). The community challenge allowed us to gain insights into modeling microbiome data and engage teams with diverse areas of expertise. Overall, the community challenge resulted in optimized models that incorporate microbiome data to provide additional biological insights into HF risk prediction.

We observed associations between incident HF occurrence and a combination of microbiome features, including alpha (intra-individual) diversity, taxonomic co-abundance networks, and individual taxonomic groups. Alpha diversity, measured with Fisher’s alpha, contributed to HF predictions in the refined models, showing an inverse association with incident HF risk. This supports a previous study which reported significantly reduced alpha) and beta (inter-individual) diversity measurements in HF patients compared to controls ^35^. The predictive co-abundance networks included several networks consisting of species (e.g *R. gnavus* (network 20), *C. bolteae* and *C. citroniae* (network 6)) that were previously reported to be positively associated with inflammatory signals ^17,26,27^. This supports a role for the gut microbiome in HF progression through inflammatory processes, a known aspect of HF pathophysiology ^36,37^. Furthermore, we observed that networks positively associated with HF are comprised of species that have been previously associated with T2D and obesity, both of which are known risk factors for HF ^37^. Thus, these established risk factors are also partially captured by the microbiome features predicting incident HF risk. However, the causal relations between the gut microbiome and established risk factors for HF warrant further research.

A large number of features is a common problem in predictive machine learning models ^38^. Thus, similar to a previous study of microbiome-based prediction of all-cause mortality ^21^, we found the approach to compress the data into a smaller number of informative characteristics by aggregating species into co-abundance networks provided a strategy for feature engineering in microbiome-based risk prediction. We also observed the network that was inversely associated with HF risk composed mostly of *Bacteroides* genera (Network 8), which have been reported to be lower in patients with cardiovascular diseases ^39^. A recent study of enterosignatures (ESs) suggested that *Bacteroides*-ES provide core functionality to the healthy gut microbiome, particularly in the Western population ^40^. Despite containing species that are associated with HF, we also observed that network 6 contains a number of bacteria, such as *Anaerostipes caccae*, a butyrate producer, that have been previously reported to be depleted in HF patients ^14^. Similarly, one network comprised mostly of Coriobacteriaceae which have been shown to have a lower abundance in patients with HF compared to controls ^35^. Overall, the findings from the co-occurrence analysis support that examining the specific interactions between previously disease-associated and health-promoting microbes can elucidate the associations within the gut microbiome in various conditions, although we expect the relevance of this strategy to vary for different traits.

We found that mean aggregations could improve model accuracy and calibration (Figure 4, Supp. Table 7) as the model performance may vary in different subsets of the data. Hence, ensemble modeling approaches provide a promising target for further enhancing the overall predictive capability and clinical relevance of ML models. By exploring different strategies for combining models and optimizing the feature selection strategies, we can potentially overcome limitations to achieve more robust predictions across diverse demographic groups.

Interestingly, we found that the microbiome-based predictions of incident HF during the 10-year follow-up period outperformed the 5- and 15-year follow-ups. While we speculate that the demographics of the population are most informative at the 10-year interval and therefore allow this increase, it remains uncertain whether this is the case. Further efforts to model incident disease with microbiome data will be necessary to understand the timing of the most informative endpoints. Likewise, focusing on the stricter definition of HF led to improved predictions, as seen in analyses of other more homogeneous phenotypes in other types of omics ^41^.

While this study offers a distinctive perspective on the prediction of incident HF risk based on population-level microbiome profiling, it must be interpreted in the context of its limitations. First, the ability of the challenge participants to optimize their models was limited since they only had access to synthetic data, which may obscure the biological link between the clinical variables and microbiome data, and access to only a limited number of submissions and computational resources. Moreover, this may have affected the types of models participants chose to use. No participants incorporated external datasets into the models. This may have limited the generalizability of the models but also represents a limitation in the public availability of suitable complementary microbiome datasets.

We used Harrell’s C-index and Hosmer-Lemeshow tests to assess model performance and calibration. While both tests are widely used, they have known limitations. Harrell’s C may not fully account for calibration. The Hosmer-Lemeshow test supports model calibration better, but it can be sensitive to the specific choice of risk groups and it may not always detect model miscalibration ^42^. Additionally, best-performing models were selected based on the real scoring set, and as a result, the model refinement performed afterward for the top models is no longer independent of the scoring set. Regarding data, the use of shallow metagenomics limited the depth of microbiome profiling and analysis. Furthermore, the taxonomic annotations pipeline choices and reference database selections could influence the results. Considering up-to-date microbial reference databases or alternative processing pipelines may impact outcomes. The study was also limited by geography and population characteristics. The generalizability of the results to demographically or geographically distinct populations will deserve further study as the study design did not consider predictors specific to demographic subgroups. Verification and extensions in independent prospective population cohorts will be important for future microbiome-based studies in cardiovascular disease. Future studies should consider an extended variety of model assessment tools for microbiome data, for example including functional measurements to gain a more comprehensive understanding of microbial communities and their activity and impact on human health status. Despite its limitations, this work contributes to establishing the groundwork for microbiome-based risk prediction of heart disease.

### Conclusions

Community challenges facilitate the comparison of alternative different modeling strategies. We have presented the outcomes of a computational modeling crowdsourced challenge to predict the risk of incident HF based on gut microbiome profiling in a prospective population cohort of Finnish adults. The results indicate that taxonomic profiles are predictive of incident HF risk and suggest an association between HF risk and specific taxonomic groups linked to inflammation. However, the incorporation of taxonomic features did not substantially improve HF risk prediction models over baseline covariates. Taken together, the results provide insights into the links between gut microbiome composition and incident HF as well as considerations for successful feature engineering and modeling strategies for prospective risk prediction in microbiome studies. Further work to generalize and interpret the models can help to elucidate the underlying causal mechanisms.

## 4. Methods

### 4.1. FINRISK data characteristics

The FINRISK study, initiated in 1972, aims to investigate risk factors associated with cardiovascular disease in Finland. The study is conducted every five years, and in 2002, participants were selected from specific areas: North Karelia, Northern Savo, Oulu, Lapland, Turku and Loimaa, Helsinki, and Vantaa (Figure 1A) ^43^. A random sample of individuals aged 24-74 years, stratified by sex and 10-year age groups, was taken from each study area. Out of the 13,498 individuals invited to participate, a total of 8,783 agreed to participate, and among them, 7,231 provided fecal samples. The data was combined with national healthcare registers in Finland, which allowed for the integration of subsequent disease diagnoses and drug prescriptions based on individual personal identity codes. The study protocol for FINRISK 2002 was approved by the Coordinating Ethics Committee of the Helsinki University Hospital District (Helsinki, Finland) with reference number 558/E3/2001, and all participants provided written informed consent. We are using the taxonomic composition characterized from the fecal samples and relevant baseline clinical variables such as BMI, age, sex, smoking status, blood pressure treatment, systolic blood pressure, and relevant disease history (diabetes, coronary heart diseases, and HF), in addition to the information of incident HF. We used a broad definition of HF based on the ICD-10 codes, I11.0, I13.0, I13.2, I50, ICD-9 codes 4029B, 428, and ICD-8 codes 42700, 42710,428,7824 in the nationwide Care Register for Health Care. In addition, HF was characterized based on three or more drug purchases with Anatomical Therapeutic Chemical drug code C03CA01, and C03EB01 in the nationwide Drug Reimbursement Register prior to baseline.

### 4.2 Metagenomic sequencing from stool samples and taxonomic profiling

Fecal sample collection, DNA extraction, and taxonomic profiling have been previously described ^21^. Briefly, the participants sent the fecal samples by mail to the laboratory of the Finnish Institute for Health and Welfare during the Finnish winter (survey period: January to March 2002). The samples were shipped overnight and stored at a temperature of -20°C upon arrival. The stool samples remained frozen until they were later transported in 2017 to the University of California San Diego for shallow shotgun metagenomics sequencing using Illumina HiSeq 4000 Systems. The DNA extraction process in this study followed the protocols of the Earth Microbiome Project, using the MagAttract PowerSoil DNA Kit. The library preparation for sequencing was conducted using a miniaturized version of the Kapa HyperPlus Illumina-compatible library prep kit. Automation systems such as the Echo 550 and Mosquito HV liquid-handling robots were used for various steps, including DNA dilution, enzymatic fragmentation, end-repair, adapter-ligation reactions, and addition of sample-specific barcoded sequences. The libraries were quantified using the PicoGreen assay and pooled in approximately equal amounts before being sequenced on an Illumina HiSeq 4000 instrument for shallow metagenomic sequencing. This protocol yielded an average read count of 900,000 reads per sample.

The sequences were subjected to quality and adapter trimming using Atropos ^44^, and host reads were eliminated by aligning them to the human genome assembly GRCh38 with Bowtie2 ^45^. Taxonomy assignment was performed using SHOGUN v1.0.5 ^46^ against a comprehensive database comprising complete bacterial, archaeal, and viral genomes available from NCBI RefSeq (as of version 82, dated May 8, 2017). A total of 5,749 taxonomic features were successfully annotated.

### 4.3. Design of the DREAM challenge

The Heart Failure Prediction: Microbiome-FINRISK DREAM Challenge was designed as a collaborative competition aimed at advancing our understanding of the relationship between gut microbiome and cardiovascular disease, particularly HF. The challenge was hosted on the Synapse platform (https://www.synapse.org/finrisk) and financially supported by the COST Action ML4Microbiome (CA18131). We provided participants access to synthetic dataset of microbiome and clinical data, as well as additional resources including a discussion forum, model submission pipeline, and leaderboards. We also provided participants with three baseline models based on Cox proportional hazards model:

1. Baseline Age-Sex which incorporates only Age and Sex information.
2. Baseline Covariates which included all relevant conventional risk factors as covariates.
3. Baseline All which builds upon Baseline Covariates by further incorporating the centered log-ratio of relative species abundance information.

Teams were required to comply with a data usage agreement, which restricted the use of data outside the Challenge and provided guidelines on ethical participation. All the participants/teams are included in the FINRISK - Heart Failure and Microbiome Challenge Community and they were required to make their code public after the challenge, provide a write-up with a detailed description of their modeling process, and participate in a post-challenge survey to collect information on method development and selected features in the model.

### 4.4. Synthetic data generation

Synthetic data provided an added layer of protection for the privacy of individuals in the FINRISK cohort. The novel approach (a separate publication is being prepared) preserves the mean and covariance structure of the original dataset, creating as many synthetic observations as in the original dataset. Cox proportional hazard models were applied to ensure that the regression associations between variables (each clinical variable and taxon) and incident HF remained. The missingness pattern for unobserved variables was recreated based on the original dataset to preserve the distribution and covariance structure. All the processes above were created independently in clinical and microbial compositional data.

### 4.5. Singularity container generation

Singularity containers were used to ensure reproducibility and to minimize the impact of differences in software versions or operating systems on the results. This provided a standardized computational environment for the server that hosted the confidential FINRISK dataset. A more detailed explanation of the Singularity container generation is provided at the challenge homepage https://www.synapse.org/#!Synapse:syn27130806/wiki/616714.

### 4.6 Model creation in the challenge environment

The synthetic dataset was divided into training (N=3615) and testing (N=1807) sets maintaining the same individual compositions in the FINRISK dataset. Participants could use the training set to build algorithms and evaluate the performance of these algorithms in the testing set. DREAM challenge participants created their prediction models based on the synthetic training dataset and submitted these for evaluation in the real FINRISK 2002 testing data set (https://www.synapse.org/finrisk). In the first phase (*submission* phase), the models were evaluated on the testing data, and the performance metrics were returned to the submitting team. In the submission phase of the challenge, a total of 35 valid submissions were received from 9 registered teams (187 registered participants, Supp. Table 2). Each team was allowed to make up to five submissions in total in order to optimize their approach. Leaderboards were available during the open phase of the challenge to offer real-time feedback and comparative performance rankings to the teams. Of these, seven teams went on to submit final model Singularity containers for evaluation during the scoring phase (Supp. Table 3). The final scoring dataset (N=1809) was not shared with participants.

At the end of the submission phase, each team selected their final model to be evaluated on the confidential scoring dataset that was not used during the submission phase. The performance of the seven models was evaluated based on their ability to predict the HF incidence in the confidential scoring dataset which was not revealed to the participants.

### 4.7. Assessment metrics

The output of each submitted model was expected to be a two-column file, with the first column containing Sample ID and the second column containing a numeric predictor for the predicted absolute HF risk for ∼15 years follow-up, with larger numbers associated with a higher probability of incident HF.

The primary metric used to assess model performance was Harrell’s C (concordance) index ^47^, a standard evaluation measure for survival models. Additionally, we used the Hosmer-Lemeshow goodness of fit method to assess model calibration. This has been designed to test whether the average of the predicted risk scores follows the observed event rate which ensures the survival models provide reliable estimates of the expected event probabilities.

To ensure a robust ranking of participants, we additionally performed 1000 bootstrap iterations of random sampling on the individual’s risk scores calculated by each model. The evaluation metrics, Harrell’s C-index and Hosmer-Lemeshow p-value, were then re-calculated to generate a distribution of evaluation scores for each submission. We used these metrics to calculate the Bayes factor, using the *computeBayesFactor* functions from the challenge scoring R package (https://github.com/Sage-Bionetworks/challengescoring/blob/develop/R/bootstrap.R) and comparing them to the top-performing model as well as to the baseline models.

### 4.8. Statistical modeling and data analysis of the two top-performing models

A schematic overview of the two top-performing models is provided in Figure 3. Notably, both top-performing teams used variations of the regularized Cox proportional hazard models, which can be depicted with the target function ^48^:

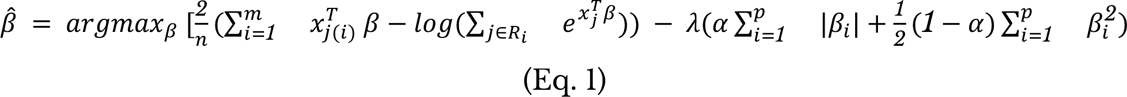

Where ! are the linear model’s regularized coefficients, 2 is the penalization term controlling the ratio between model goodness-of-fit (first term) and the coefficient shrinkage toward zero (second term) searched in a grid with cross-validation, and 3 is the ratio of LASSO-regularization (3= 1, L1 norm), Ridge Regression (3 = 0, L2 norm), and their Elastic Net (EN) mixture (0 < 3 < 1). For DFH team, Eq. 1 provided the regularized final model coefficients, while for SB2 Eq. 1, with 3=1 was only used for feature selection after which non-regularized Cox model fit provided the model coefficients.

To reduce the dimensionality of the data, SB2 team used a regularized Cox proportional hazard model (based on LASSO penalty) to select the most relevant features. The unpenalized features were used to build a Cox proportional hazards model. Prior to conducting analyses, the taxonomic features were aggregated at the species level, and features that were not associated with known species were removed. Four types of features were then calculated for each volunteer based on the microbiome data. (i) Firmicutes-Bacteroidetes ratio, as this ratio is often associated with dysbiosis in earlier literature ^12,49^. (ii) the relative abundance of all phyla. (iii) various diversity indices including Shannon, observed richness, Chao1, Simpson, inverse Simpson, and Fisher’s alpha indices. (iv) GSVA scores for each co-abundance network^50^. The species that did not have any significant associations with other species based on 50 bootstrap iterations (p < 0.01) using data from all individuals in the training dataset were excluded.

The Louvain method was used as an unsupervised two-step algorithm for community detection with a resolution value of four ^51^. There were 25 distinct networks of co-abundant species identified after the process. The overall abundance of each such co-abundance network was then summarized with microbial set variation analysis using GSVA ^52^. This reduced the dimensionality of the data by agglomerating strongly co-varying species. The co-abundance networks were obtained based on the training set, and subsequently also used with the testing and scoring sets. A total of 50 features were thus obtained from the combination of clinical and microbiome data.

Finally, to further reduce dimensionality, simplify the model, and mitigate overfitting, a Cox regularized regression with LASSO feature selection was employed using the biospear package ^53^. The weight of Firmicutes-Bacteroidetes ratio as a proxy for dysbiosis and clinical covariates including age, BMI, systolic blood pressure, non-HDL cholesterol were fixed and not optimised by LASSO analysis. The features selected by LASSO were then used as input variables in a Cox model. The inverse survival probability was calculated with the *survfit* function from the survival R package ^54^ to obtain risk score for each individual in the scoring dataset.

The DFH team took an overall similar approach to the SB2 team, leveraging regularized regression (specifically, Cox regularized regression) to generate informative modules from volunteer microbiome data in conjunction with clinical metadata, complemented by literature-derived findings to guide interpretability^48^.

The approach consisted of nested feature selection via Elastic Net (EN, *ɑ* = 0.5 in Eq. 1) penalization for Cox proportional hazards models. Modules consisting of connected candidate features were manually curated. Each module was independently subjected to EN regularization in order to prune the module to its most informative predictors, with the feature selection conducted using the first conservative regularization parameter 6 within 1 standard error of the local optimum in 10-fold cross-validation (CV) (Eq.1). Second, these pruned modules were brought together and whole modules were eliminated using the same CV strategy. Finally, these two steps were run using a multi-seeded CV, after which the predictions produced over the seeds were averaged over to reduce effects from random binning in CV. These modules submitted in the final challenge model were as follows: i) Two clinical submodules, one with a focus on age and sex, and the other with a focus on more complex interactions and self-derived variables such as an ad hoc disease burden estimate; ii); Alpha and beta diversity indices for within-sample and between-sample variation; iii) A comprehensive list of relative abundances of taxa in varying levels, coupled with two modules focusing on literature-derived findings; iv) Family and phylum level relative abundances coupled with binarized variables for age, sex, and obesity. Further, to expand to non-linear interactions between variables, transformations (such as z-score, square root, power of 2, or x log(x)) were used, along with taking the product of multiplying candidate features to assess their combined effect after transformations. The final submission included modules presented in teams Wiki (https://www.synapse.org/#!Synapse:syn50613972/wiki/620396). Multiple varying transformations were used to introduce non-linearity into the model components.

### 4.8. Model refinement during the post-challenge phase

Due to participants’ limited access to the real dataset, we also worked with the top two performing teams to further improve the model performance and calibrations for the final submitted models after the challenge formally concluded. The ensemble model was created by simple mean aggregations of the individual risk score of an increasing number and all possible combinations from all submissions (Figure 4).

### 4.9. Model evaluation in stricter HF definition and 5-, 10-year follow-up

We also tested the models using a stricter definition of HF (N cases=288) ^34^. We further examined model performance with a restricted follow-up time of 5 and 10 years, in addition to the 15-year interval used in the challenge. For this analysis, we focused only on the baseline and the refined models in the post-challenge phase. The evaluation criteria remained consistent with those applied during the challenge, which are Harrel’s C-index and the Hosmer-Lemeshow p-value.

## Data Availability

Code and model descriptions during the final phase of the challenge is available online, which may be accessed by the hyperlinks listed in Supp. Table 3. The synthetic and real data used in the challenge can be obtained upon a reasonable request from THL Biobank, https://thl.fi/en/web/thl-biobank/for-researchers/ due to sensitive health information of individuals.

## FINRISK Microbiome Dream Challenge Community SB2

1. José Liñares-Blanco^1,2,3^
2. Pedro Carmona-Saez^1,2^
3. Samuel Pérez-Fernández^1,2^
4. Marina Vargas^1,2^
5. Iván Ellson-Lancho^1^
6. Juan A. Villatoro^1,2^
7. Raúl López-Domínguez^1,2^
8. Jordi Martorell-Marugan^1,4^
9. Daniel Toro-Domínguez^1^
10. Adrián García-Moreno^1^

## DenverFINRISKHacky

1. Teemu D. Laajala^5,6^
2. Lily Elizabeth Feldman^5^
3. Varsha Sreekanth^5^
4. Michael Orman^5^

## Yuanfang Guan Lab Team

1. Hanrui Zhang^7^
2. Yuanfang Guan^7^
3. Yiyang Nan^7^

## Metformin-121

1. Chih-Han Huang^8^
2. Tsai-Min Chen^9,10^
3. Hsuan-Kai Wang: Independent Researcher
4. Edward S.C. Shih^11^
5. Kuei-Lin Huang^12^
6. Chih-Hsun Wu^13^
7. Sz-Hau Chen^14^
8. Jhih-Yu Chen^15^

## Individual participant

1. Tristan Fauvel^16^,

## UTK-Bioinformatics_FINRISK

1. Ashley Babjac
2. Nahian Tahmin^17^
3. Anahita Khojandi
4. Scott Emrich

## Pteam

1. Renato Giliberti^18^

## Affiliation

1. GENYO. Centre for Genomics and Oncological Research: Pfizer, University of Granada, Andalusian Regional Government, PTS Granada, Avenida de la Ilustración 114, 18016, Granada, Spain
2. Department of Statistics and Operations Research, University of Granada, Spain
3. Heidelberg University, Faculty of Medicine, and Heidelberg University Hospital, Institute for Computational Biomedicine, Bioquant, Heidelberg, Germany.
4. Fundación para la Investigación Biosanitaria de Andalucía Oriental-Alejandro Otero (FIBAO), 18012, Granada, Spain.
5. Department of Pharmacology, University of Colorado Anschutz Medical Campus, Aurora, CO 80045, USA.
6. Department of Mathematics and Statistics, Faculty of Science, University of Turku, Finland
7. University of Michigan: Ann Arbor, MI, US
8. ANIWARE, Taipei, Taiwan
9. Graduate Program of Data Science, National Taiwan University and Academia Sinica, Taipei, Taiwan,
10. Research Center for Information Technology Innovation, Academia Sinica, Taipei, Taiwan
11. Institute of Biomedical Sciences, Academia Sinica, Taipei, Taiwan
12. Chung Shan Medical University Hospital, Taichung, Taiwan
13. Artificial Intelligence and E-learning Center, National Chengchi University, Taipei, Taiwan
14. Development Center for Biotechnology, Taipei, Taiwan
15. Graduate Institute of Biomedical Electronics and Bioinformatics, National Taiwan University, Taipei, Taiwan
16. Quinten Health, Paris, France
17. University of Tennessee at Knoxvilee
18. University of Naples Federico II, Napoli NA, Italy

## ML4Microbiome Community

**The ML4Microbiome Community,** under the **COST Action CA18131** (**Statistical and machine learning techniques in human microbiome studies)**, is part of COST Actions, a research network funded for four years. It requires participation from at least seven different COST Full Members or Cooperating Members. These Actions use various networking tools to support research coordination and capacity building, as specified in each Action’s Memorandum of Understanding. COST, a funding agency for research and innovation networks in Europe, provides the funding for these Actions. **ML4Microbiome** introduced the idea for the community challenge, and some of its members were involved in organizing the challenge as part of the realizations of Working Group 2. You can find more information about the community here: https://www.ml4microbiome.eu/.

## Acknowledgements

This article is based upon work from COST Action ML4Microbiome “Statistical and machine learning techniques in human microbiome studies”, CA18131, supported by COST (European Cooperation in Science and Technology), www.cost.eu. Thanks to Sage Bionetwork/ DREAM for assistance in organizing and hosting this DREAM Challenge. The authors thank all participants in the FINRISK 2002 study and Tara Schwartz for assistance with laboratory work. We would like to also thank Attila Gabor and Jovan Tanevski for their valuable and insightful discussions.

## Statement of Funding

This work was supported by COST (European Cooperation in Science and Technology) and Informatics4Life, a joint initiative founded by the Klaus-Tschira Foundation. P.P.E. is funded by Turku Collegium for Science, Medicine, and Technology (TCSMT). R.T.L. & E.K. are funded by Informatics4life. T.D.L. was supported by the Finnish Cultural Foundation. L.E.F was supported by the Pharmacology Training Grant (T32 GM007635) through the University of Colorado Anschutz Medical Campus. T.N. is funded by the Sigrid Jusélius Foundation, the Finnish Foundation for Cardiovascular Research, and the Research Council of Finland (grants 321531 and 354447). A.S.H. was supported by the Research Council of Finland (grant 321356). J.S.-R. has received funding from GSK, Pfizer and Sanofi, and fees/honoraria from Travere Therapeutics, Stadapharm, Astex, Pfizer and Grunenthal. L.L. was supported by Research Council of Finland (grant 330887).

## Conflict of Interest

Illumina, Inc., and Janssen Pharmaceutica provided additional support by sponsoring the Center for Microbiome Innovation at the University of California San Diego. T.N. has received honoraria for speaking engagements from Servier and AstraZeneca. V.S. has had research collaboration with Bayer AG, unrelated to this study. J.S.-R. has received funding from GSK, Pfizer and Sanofi, and fees/honoraria from Travere Therapeutics, Stadapharm, Astex, Pfizer and Grunenthal. M.I. is a trustee of the Public Health Genomics (PHG) Foundation, a member of the Scientific Advisory Board of Open Targets, and has a research collaboration with AstraZeneca unrelated to this study. R.K. is a cofounder of Micronoma and Biota, holding stock for Gencirq, Cybele, Biomesense, Micronoma, and Biota, serve as a member of the Scientific Advisory Board in Gencirq, DayTwo, Biomesense, and Micronoma and serve as consultant for DayTwo, Cybele, and Biomesense.

## Authors contributions

P.P.E., E.K. analyzed the data, developed models for the challenge, organized the challenge, and wrote the manuscript. J.L.B., T.D.L., L.E.F., and P.C.S. developed models for the challenge, provided feedback on model performance, contributed to data analysis, and helped write the manuscript. R.S. organized the challenge and helped write the manuscript. M.J.C., R.J.B., D.C.G., and S.M. contributed to the challenge initiation and design. J.A. and V.C. gave technical support for the challenge organization via Sage Bionetworks. M.I., P.J., J.H.S., and H.C.F. provided intellectual input on study goals and design and contributed to manuscript preparation. R.K. acquired shotgun metagenomics data. V.S. and T.N. contributed to patient recruitment and the collection of biomaterials and clinical data. A.S.H. conceived of the study, designed the study, created the synthetic data, and contributed to data analysis. M.J.C., R.J.B., D.C.G., J.S.R., R.T.L., and L.L. conceived the study. J.S.R., R.T.L., and L.L. designed the study, organized the challenge, helped interpret the data, and helped write the manuscript. All authors reviewed, edited, and approved the final version of the manuscript.

## Notes

### Author Declarations

The Coordinating Ethics Committee of the Helsinki University Hospital District approved the study protocol for FINRISK 2002 (Ref. 558/E3/2001)

