## Supplementary material for "Microbiome-based risk prediction in incident heart failure: a community challenge": Supplementary_materials.pdf

### Supplementary Figure & Tables

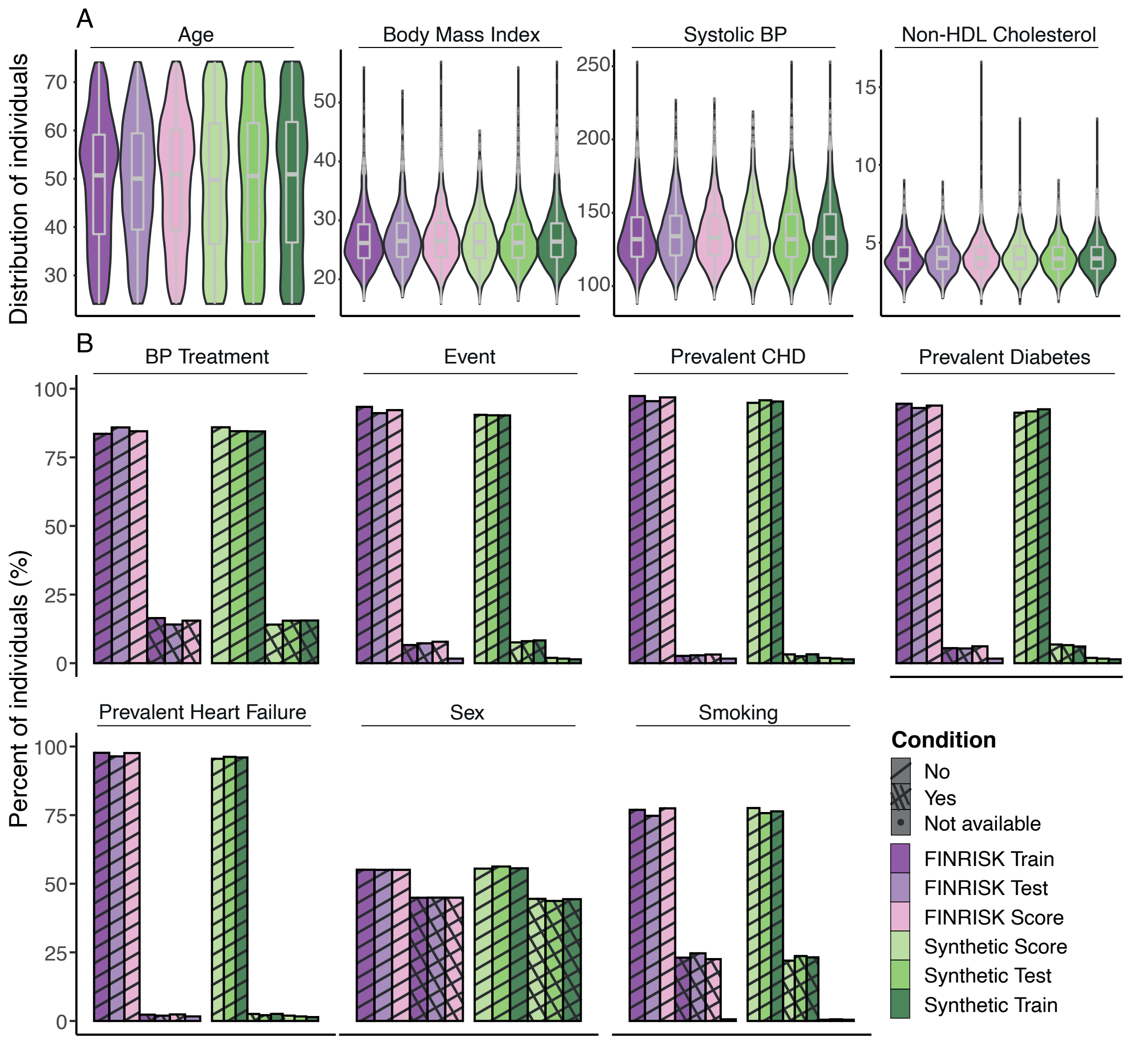

**Supplementary Figure 1. Comparison of Synthetic and FINRISK Datasets for (a) Continuous and (b) Categorical Covariates.**

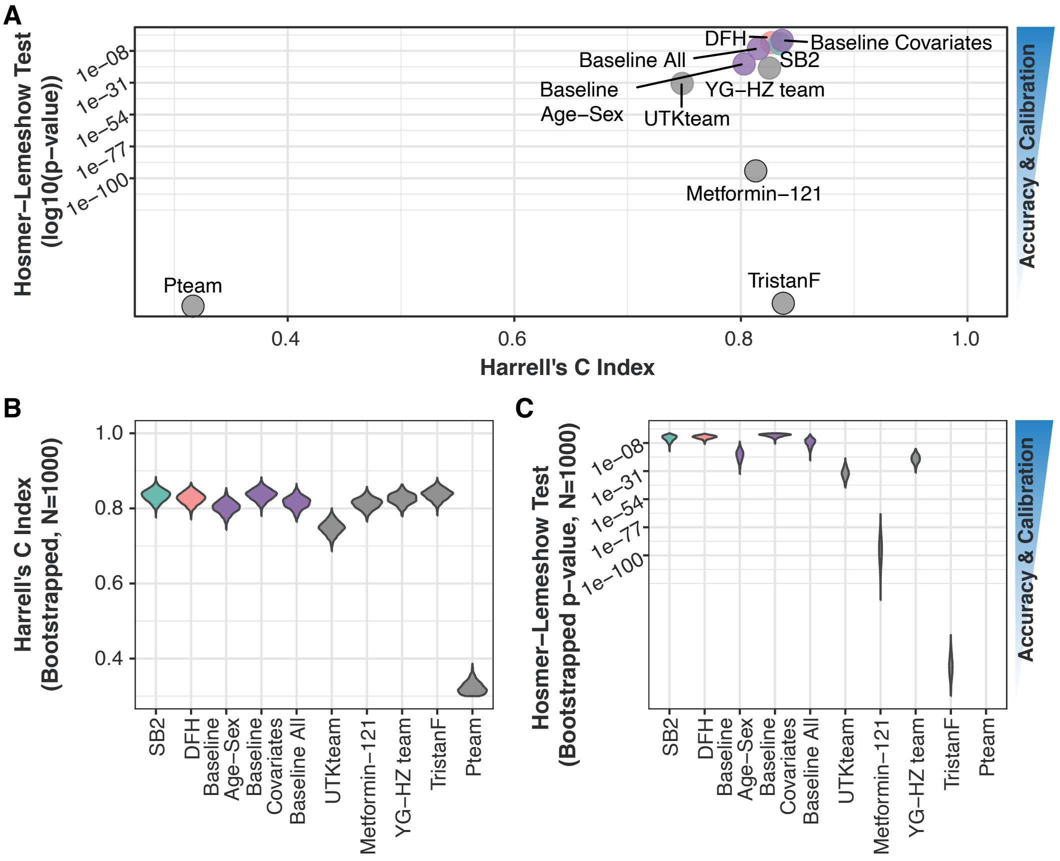

**Supplementary Figure 2. Harrell's C (model discrimination measure) and Hosmer-Lemeshow test (model calibration measure) results. (A.)** Harrell's C-index and (**B.**) Hosmer-Lemeshow p-value were obtained for the investigated models, including the three baseline models provided by the organizers in the scoring phase. **C-D.** Harrell's C-index and Hosmer-Lemeshow empirical p-value on 1000 bootstrapped iterations for all the models.

**A. Network 1**

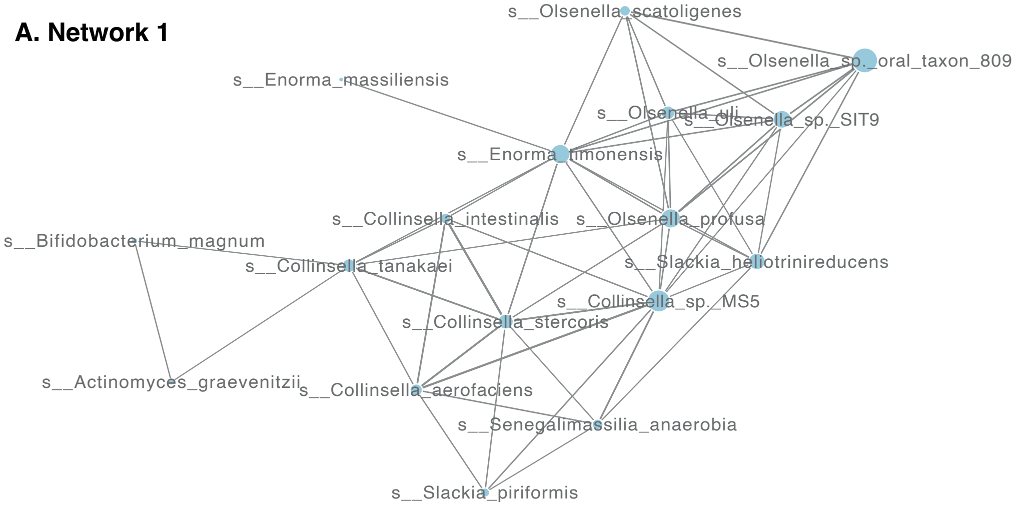

**B. Network 2**

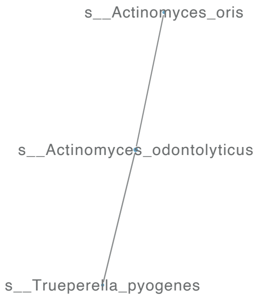

**C. Network 5**

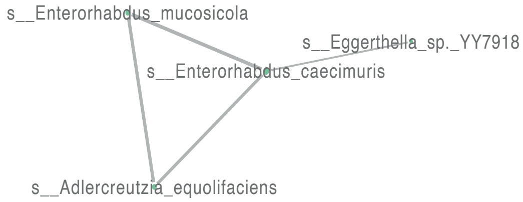

**D. Network 6**

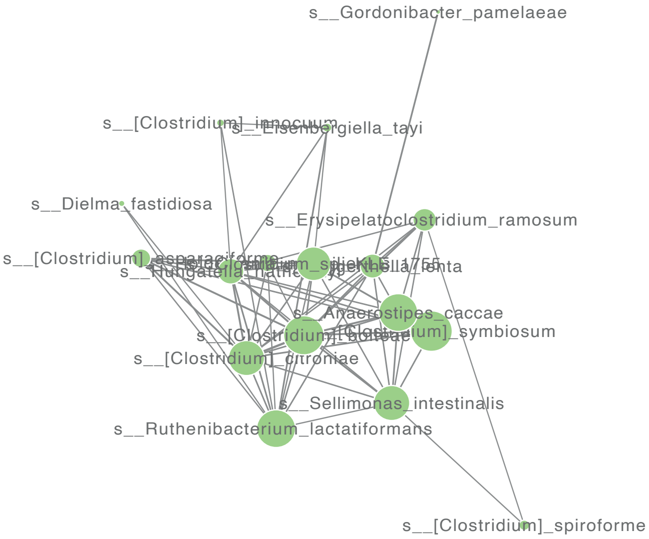

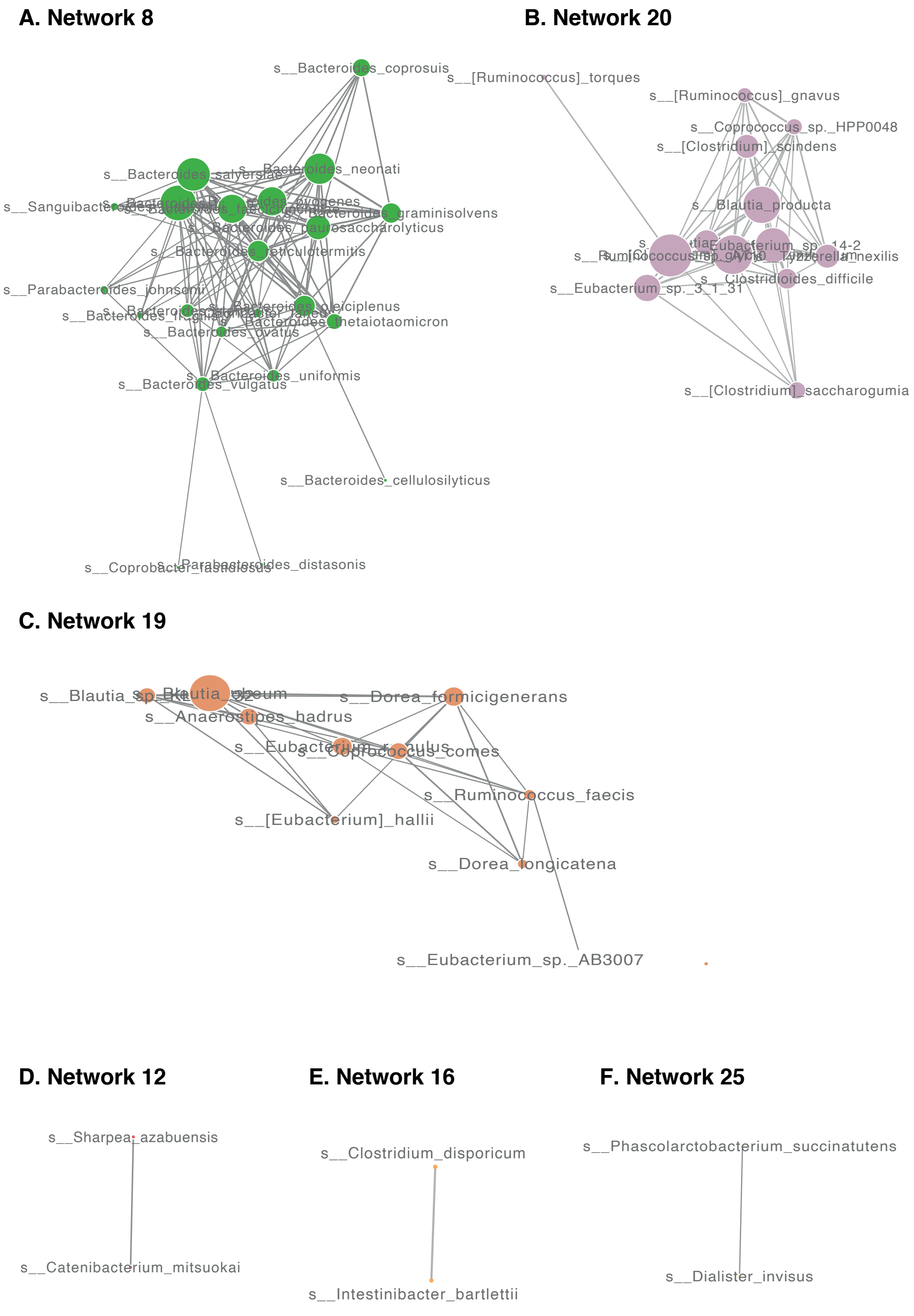

**Supplementary Figure 3-4. Taxonomic co-abundance networks from SB2 team refined model.**

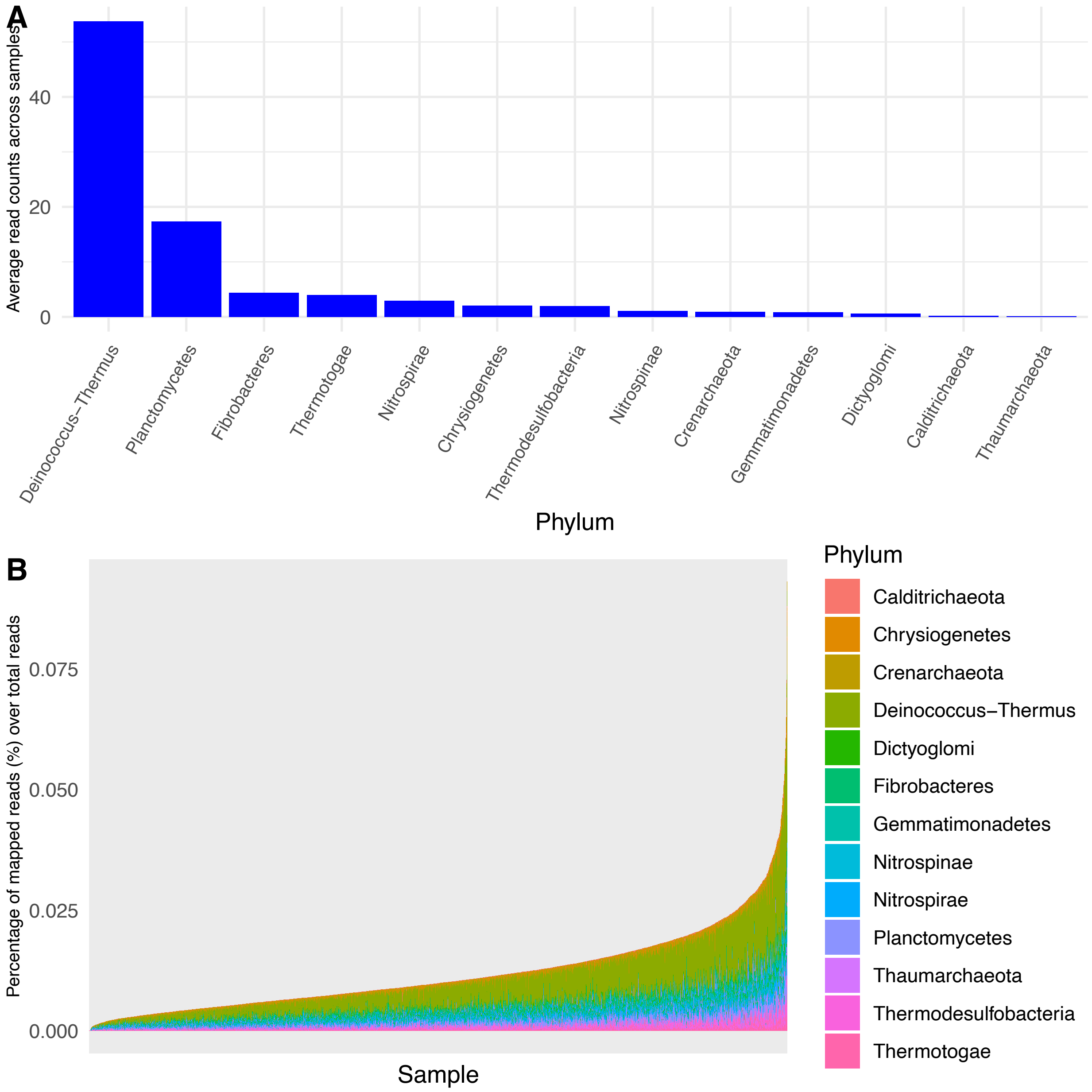

**Supplementary Figure 5. Reads count distributions in the infrequently reported phyla.** (A.) Average read counts across samples per infrequently reported phyla. (B.) The percentage of read counts assigned to infrequently reported o phyla in each sample.

**Supplementary Table 1. Comparison of National FINRISK 2002 vs Synthetic dataset**

| Variable | Synthetic Dataset | FINRISK 2002 Dataset | P-value |
|---|---|---|---|
| Number | 7231 | 7231 | |
| Richness | 195.22 (37.57) | 189.46 (36.81) | 1,10E-25 |
| exp(Shannon) | 32.2 (11.59) | 31.37 (11.43) | 2,80E-05 |
| inv(Simpson) | 15.1 (6.95) | 14.83 (6.89) | 0,017 |
| evenness (q1) | 0.16 (0.04) | 0.16 (0.04) | 0,24 |
| Age | 49.35 (14.83) | 49.47 (12.97) | 0,9 |
| Body Mass Index | 26.97 (4.7) | 27.01 (4.69) | 0,43 |
| Systolic Blood Pressure | 136.29 (22.09) | 135.81 (20.33) | 0,92 |
| NonHDL Cholesterol | 4.09 (1.1) | 4.09 (1.09) | 0,92 |
| Event time | 13.76 (5.58) | 13.46 (4.27) | 0 |
| Sex | 3199 (44.2%) | 3248 (44.9%) | 0,42 |
| Smoking | 1663 (23%) | 1687 (23.3%) | 0,65 |
| Blood Pressure Treatment | 1096 (15.2%) | 1129 (15.6%) | 0,46 |
| Prevalent Diabetes | 462 (6.4%) | 405 (5.6%) | 0,05 |
| Prevalent Coronary Heart Disease | 221 (3.1%) | 205 (2.8%) | 0,46 |
| Prevalent Heart Failure | 177 (2.4%) | 162 (2.2%) | 0,38 |
| Event | 582 (8%) | 559 (7.73%) | 0,0052 |

Data are presented as n (%) (n of participants in indicated category and percentage of total) or mean (SD).
*Mann-Whitney U test was used for numeric data; Fisher exact test was used for categorical data.

**Supplementary Table 2 . Valid submission during the first phase (submission phase)**

| ID | Entity ID | Submitter ID | Harrell's C index (Test Data) | Hosmer-Lemeshow P-value (Test Data) |
|---|---|---|---|---|
| 9730535 | syn50690974 | Pteam | 0.964662114389025 | 0.0027391484623173 |
| 9730509 | syn50679121 | TristanF | 0.85550874385106 | 3.68376485511634e-165 |
| 9730516 | syn50680052 | TristanF | 0.85550874385106 | 3.6837648551155e-165 |
| 9730289 | syn50619085 | Metformin-121 | 0.855499329206581 | 0.0000805580909573684 |
| 9730539 | syn50691116 | RH_FP | 0.855461670628663 | 0.124062184456823 |
| 9730540 | syn50691121 | RH_FP | 0.855461670628663 | 0.017109223667752 |
| 9730537 | syn50691084 | Metformin-121 | 0.854619059947749 | 0.0003183735189758 |
| 9730494 | syn50680115 | SB2 | 0.853639936921882 | 1.43329135289717e-172 |
| 9729604 | syn48184532 | SB2 | 0.852905594652482 | 1.01380073567315e-302 |
| 9730252 | syn50559327 | Metformin-121 | 0.852505472262104 | 0.626600030695395 |
| 9730529 | syn50685722 | Metformin-121 | 0.851695812836868 | 0.176194374513841 |
| 9730270 | syn50614047 | DFH | 0.847647515710688 | 4.03783956084775e-73 |
| 9730210 | syn50549160 | YG-HZ team | 0.845571586602961 | 1.4586210486304399e-18 |
| 9730448 | syn50679023 | SB2 | 0.845270317979617 | 0 |
| 9730512 | syn50679982 | YG-HZ team | 0.844427707298703 | 5.2351323279863495e-36 |
| 9730510 | syn50680913 | Metformin-121 | 0.843312071927884 | 4.3973620031984897e-11 |
| 9730513 | syn50679981 | YG-HZ team | 0.842540071080566 | 6.051285277708881e-44 |
| 9728266 | syn42923449 | TristanF | 0.835803892955492 | 6.45936231117481e-10 |
| 9730523 | syn50680056 | TristanF | 0.835504977993269 | 0.0000000111636762077687 |
| 9730495 | syn50680126 | DFH | 0.834514086661802 | 2.48841432923713e-69 |
| 9730532 | syn50688995 | DFH | 0.833713841881046 | 0 |
| 9730530 | syn50558657 | TristanF | 0.83253465765999 | 1.5425277407005702e-14 |
| 9730451 | syn50679131 | DFH | 0.831765010473792 | 9.423486133224709e-147 |
| 9730031 | syn50548019 | YG-HZ team | 0.819149386871278 | 1.22707483366914e-42 |
| 9730534 | syn50689118 | DFH | 0.8165462376727 | 0 |
| 9729772 | syn49501945 | SB2 | 0.800574293313249 | 6.29334377950434e-31 |
| 9730514 | syn50680923 | SB2 | 0.790493562736837 | 1.34923083061534e-42 |
| 9730533 | syn50688910 | Team 2016 | 0.74970935336462 | 6.36823787800633e-13 |
| 9729151 | syn44768675 | A_lab | 0.748710477989152 | 1.87818159351935e-48 |
| 9729620 | syn48550025 | A_lab | 0.693869609987805 | 0.0024241535415921 |
| 9729767 | syn49498137 | A_lab | 0.693869609987805 | 0.0024497170647116 |
| 9730211 | syn50549194 | YG-HZ team | 0.57297997034387 | 4.03322899800313e-108 |
| 9730515 | syn50680925 | RH_FP | 0.548334425092478 | 0 |
| 9730511 | syn50680919 | RH_FP | 0.532308374742227 | 0 |
| 9730517 | syn50680926 | RH_FP | 0.511026551023894 | 0 |

| Harrell's C index (Train Data) | Hosmer-Lemeshow P-value (Train Data) |
|---|---|
| 0.981833804173773 | 1.84349159550014e-30 |
| 0.848888859958601 | 0 |
| 0.84892401025868 | 0 |
| 0.898446877481676 | 0.000000215534923443216 |
| 0.848759975524976 | 0.588315584458332 |
| 0.848759975524976 | 0.572750024914225 |
| 0.925641492976449 | 9.526428969917151e-15 |
| 0.848759975524976 | 0.57275002491423 |
| 0.939646934763647 | 4.52696817617866e-32 |
| 0.847647515710688 | 4.03783956084775e-73 |
| 0.845571586602961 | 1.4586210486304399e-18 |
| 0.845270317979617 | 0 |
| 0.849848332964472 | 6.51022447407392e-107 |
| 0.868270995794983 | 0.0075261755632396 |
| 0.95908895629646 | 5.54977129812122e-102 |
| 0.843551872729876 | 1.2203464599551299e-22 |
| 0.865749287230026 | 1.76571653675929e-22 |
| 0.826677775897309 | 1.32289310167985e-186 |
| 0.829761889263536 | 0 |
| 0.841814536601877 | 1.8366684530546698e-28 |
| 0.825161105542031 | 3.1664146994484e-292 |
| 0.825166963925377 | 1.40392788716668e-87 |
| 0.829761889263536 | 0 |
| 0.875553617226251 | 1.45991493228694e-29 |
| 0.790493562736837 | 1.34923083061534e-42 |
| 0.984757788395194 | 1.72472656260911e-112 |
| 0.991983127855962 | 1.06237179217406e-26 |
| 0.991983127855962 | 1.06237179217406e-26 |
| 0.987424003749365 | 3.0136537459105004e-26 |
| 0.510314660278859 | 0 |
| 0.532308374742227 | 0 |
| 0.511026551023894 | 0 |

**Supplementary Table 3. Results and model descriptions for the final phase**

| ID | Teams | Abbreviation* | Model description | Method category | Harrel's C index | Hosmer-Lemeshow P- | Writeup |
|---|---|---|---|---|---|---|---|
| **9731637** | **SB2** | **SB2** | **Cox Proportional-Hazard** | **Regression based** | **0,8344** | **1,80E-03** | https://www.synapse.org/#!Synapse:syn47950082 |
| **9731625** | **DenverFINRISKHacky** | **DFH** | **Cox Proportional-Hazard** | **Regression based** | **0,8271** | **9,20E-03** | https://www.synapse.org/#!Synapse:syn50613972 |
| 9731636 | TristanF | TristanF | Cox Proportional-Hazard | Regression based | 0,8374 | 4,99E-191 | https://www.synapse.org/#!Synapse:syn41016074 |
| 9731490 | Yuanfang Guan and Hanrui Zhang | YG-HZ team | Guanrank, to model the survival risk for heart failure | kernel & tree-based models | 0,8252 | 4,10E-21 | https://www.synapse.org/#!Synapse:syn43580957 |
| 9731454 | Metformin-121 | Metformin-121 | XGBoost & Light Gradient Boosting Machine (LightGBM) | Regression based | 0,8131 | 2,06E-95 | https://www.synapse.org/#!Synapse:syn45414006 |
| 9731713 | UTK-Bioinformatics_FINRISK | UTKteam | Gradient Boosted Trees built with a reduced latent space | Tree-based | 0,7482 | 2,90E-32 | https://www.synapse.org/#!Synapse:syn46735476 |
| 9731666 | Pteam | Pteam | Random forest regression implemented in MetAML | Regression based | 0,3164 | 0,00E+00 | https://www.synapse.org/#!Synapse:syn50680017 |

**Top 2 performance model**
* shorter name use in manuscript and figures for clarity

Supplementary Table 4. Features coefficient and Hazzard ratio of the top 2 performing teams and post-challenge refined model

| Features | Coefficient | Model | HR | 95% CI | P-value | P-adjusted(FDR) |
|---|---|---|---|---|---|---|
| Age | 0,0990 | SB2 Final | 1,10 | (1.1e+00-1.1e+00) | 2,20E-35 | 6,40E-34 |
| Body Mass Index | 0,0820 | SB2 Final | 1,10 | (1.1e+00-1.1e+00) | 7,40E-10 | 1,10E-08 |
| Systolic Blood Pressure | 0,0087 | SB2 Final | 1,00 | (1.0e+00-1.0e+00) | 4,30E-03 | 1,40E-02 |
| NonHDL Cholesterol | -0,0550 | SB2 Final | 0,95 | (8.4e-01-1.1e+00) | 3,90E-01 | 4,90E-01 |
| Sex | 0,2700 | SB2 Final | 1,30 | (1.0e+00-1.7e+00) | 3,30E-02 | 7,30E-02 |
| Dysbiosis | -0,0017 | SB2 Final | 1,00 | (9.9e-01-1.0e+00) | 4,40E-01 | 5,30E-01 |
| Smoking | 0,5100 | SB2 Final | 1,70 | (1.2e+00-2.3e+00) | 2,20E-03 | 1,10E-02 |
| Blood Pressure Treatment | 0,4200 | SB2 Final | 1,50 | (1.2e+00-2.0e+00) | 1,60E-03 | 9,20E-03 |
| Prevalent Diabetes | 0,7900 | SB2 Final | 2,20 | (1.6e+00-3.1e+00) | 3,50E-06 | 3,40E-05 |
| Prevalent CHD | 0,7000 | SB2 Final | 2,00 | (1.2e+00-3.2e+00) | 4,00E-03 | 1,40E-02 |
| p__Crenarchaeota | -79000,0000 | SB2 Final | 0,00 | (0.0e+00-0.0e+00) | 6,50E-03 | 1,80E-02 |
| p__Bacteroidetes | -0,3100 | SB2 Final | 0,73 | (2.2e-01-2.4e+00) | 6,10E-01 | 6,10E-01 |
| p__Chlamydiae | -2000,0000 | SB2 Final | 0,00 | (0.0e+00- Inf) | 1,70E-01 | 2,60E-01 |
| p__Chrysiogenetes | 25000,0000 | SB2 Final | Inf | ( Inf- Inf) | 8,50E-03 | 2,10E-02 |
| p__Ignavibacteriae | 4700,0000 | SB2 Final | Inf | (0.0e+00- Inf) | 5,50E-01 | 5,70E-01 |
| p__Lentisphaerae | -50000,0000 | SB2 Final | 0,00 | (0.0e+00- Inf) | 1,20E-01 | 1,90E-01 |
| p__Nitrospirae | -27000,0000 | SB2 Final | 0,00 | (0.0e+00-4.2e+242) | 5,50E-02 | 1,10E-01 |
| p__Proteobacteria | 0,4300 | SB2 Final | 1,50 | (3.8e-01-6.3e+00) | 5,50E-01 | 5,70E-01 |
| p__Spirochaetes | -14,0000 | SB2 Final | 0,00 | (2.8e-24-3.1e+11) | 5,00E-01 | 5,60E-01 |
| p__Synergistetes | 88,0000 | SB2 Final | Inf | (4.8e-06-5.4e+81) | 8,50E-02 | 1,60E-01 |
| p__Verrucomicrobia | -5,1000 | SB2 Final | 0,01 | (1.1e-05-3.8e+00) | 1,20E-01 | 1,90E-01 |
| Network 1 | 0,3300 | SB2 Final | 1,40 | (9.5e-01-2.0e+00) | 8,70E-02 | 1,60E-01 |
| Network 2 | -0,3300 | SB2 Final | 0,72 | (5.6e-01-9.1e-01) | 6,80E-03 | 1,80E-02 |
| Network 6 | 0,5200 | SB2 Final | 1,70 | (1.2e+00-2.4e+00) | 3,90E-03 | 1,40E-02 |
| Network 7 | -0,1300 | SB2 Final | 0,88 | (6.3e-01-1.2e+00) | 4,60E-01 | 5,30E-01 |
| Network 8 | -0,1900 | SB2 Final | 0,83 | (5.4e-01-1.3e+00) | 3,80E-01 | 4,90E-01 |
| Network 12 | 0,1600 | SB2 Final | 1,20 | (9.2e-01-1.5e+00) | 2,10E-01 | 3,00E-01 |
| Network 19 | -0,5100 | SB2 Final | 0,60 | (4.4e-01-8.2e-01) | 1,50E-03 | 9,20E-03 |
| Network 25 | 0,1500 | SB2 Final | 1,20 | (9.1e-01-1.5e+00) | 2,30E-01 | 3,20E-01 |
| Age | 0,0989 | SB2 Phylum removed | 1,10 | (1.09-1.12) | 5,00E-36 | 1,00E-34 |
| Body Mass Index | 0,0857 | SB2 Phylum removed | 1,09 | (1.06-1.12) | 1,40E-10 | 1,40E-09 |
| Systolic Blood Pressure | 0,0086 | SB2 Phylum removed | 1,01 | (1.00-1.01) | 4,70E-03 | 1,20E-02 |
| NonHDL Cholesterol | -0,0598 | SB2 Phylum removed | 0,94 | (0.83-1.07) | 3,50E-01 | 4,10E-01 |
| Sex | 0,2878 | SB2 Phylum removed | 1,33 | (1.04-1.71) | 2,50E-02 | 4,90E-02 |
| Dysbiosis | -0,0013 | SB2 Phylum removed | 1,00 | (0.99-1.00) | 5,20E-01 | 5,50E-01 |
| Smoking | 0,4766 | SB2 Phylum removed | 1,61 | (1.16-2.23) | 3,90E-03 | 1,20E-02 |
| Blood Pressure Treatment | 0,3953 | SB2 Phylum removed | 1,48 | (1.15-1.92) | 2,80E-03 | 1,10E-02 |
| Prevalent Diabetes | 0,7224 | SB2 Phylum removed | 2,06 | (1.48-2.87) | 2,10E-05 | 1,40E-04 |
| PrevalentCHD | 0,6804 | SB2 Phylum removed | 1,97 | (1.23-3.17) | 4,80E-03 | 1,20E-02 |
| Fisher index | -0,0031 | SB2 Phylum removed | 1,00 | (0.99-1.00) | 3,30E-02 | 6,00E-02 |
| Network 1 | 0,3715 | SB2 Phylum removed | 1,45 | (0.99-2.13) | 5,80E-02 | 8,90E-02 |
| Network 2 | -0,3045 | SB2 Phylum removed | 0,74 | (0.58-0.94) | 1,20E-02 | 2,70E-02 |
| Network 6 | 0,4003 | SB2 Phylum removed | 1,49 | (0.99-2.24) | 5,30E-02 | 8,90E-02 |
| Network 7 | -0,1631 | SB2 Phylum removed | 0,85 | (0.61-1.18) | 3,30E-01 | 4,10E-01 |
| Network 8 | -0,3010 | SB2 Phylum removed | 0,74 | (0.50-1.09) | 1,20E-01 | 1,80E-01 |
| Network 12 | 0,1417 | SB2 Phylum removed | 1,15 | (0.91-1.47) | 2,50E-01 | 3,30E-01 |
| Network 19 | -0,6044 | SB2 Phylum removed | 0,55 | (0.40-0.76) | 2,60E-04 | 1,30E-03 |
| Network 20 | 0,2042 | SB2 Phylum removed | 1,23 | (0.79-1.91) | 3,70E-01 | 4,10E-01 |
| Network 25 | 0,0760 | SB2 Phylum removed | 1,08 | (0.83-1.39) | 5,60E-01 | 5,60E-01 |
| Age | 0,0992 | SB2 ageFix | 1,10 | (1.09-1.12) | 5,70E-36 | 1,30E-34 |
| Body Mass Index | 0,0851 | SB2 ageFix | 1,09 | (1.06-1.12) | 2,00E-10 | 2,30E-09 |
| Smoking | 0,4833 | SB2 ageFix | 1,62 | (1.17-2.25) | 3,60E-03 | 1,40E-02 |
| Blood Pressure Treatment | 0,3972 | SB2 ageFix | 1,49 | (1.15-1.93) | 2,70E-03 | 1,20E-02 |
| Prevalent Diabetes | 0,6910 | SB2 ageFix | 2,00 | (1.42-2.80) | 6,00E-05 | 4,60E-04 |
| PrevalentCHD | 0,6927 | SB2 ageFix | 2,00 | (1.24-3.22) | 4,30E-03 | 1,40E-02 |
| Systolic Blood Pressure | 0,0086 | SB2 ageFix | 1,01 | (1.00-1.01) | 5,10E-03 | 1,50E-02 |
| NonHDL Cholesterol | -0,0531 | SB2 ageFix | 0,95 | (0.84-1.07) | 4,10E-01 | 4,90E-01 |
| Sex | 0,3092 | SB2 ageFix | 1,36 | (1.06-1.75) | 1,60E-02 | 3,80E-02 |
| Fisher | -0,0030 | SB2 ageFix | 1,00 | (0.99-1.00) | 4,40E-02 | 9,20E-02 |
| Network 1 | 0,3576 | SB2 ageFix | 1,43 | (0.97-2.11) | 7,00E-02 | 1,30E-01 |
| Network 2 | -0,3158 | SB2 ageFix | 0,73 | (0.57-0.93) | 1,00E-02 | 2,50E-02 |
| Network 6 | 0,3440 | SB2 ageFix | 1,41 | (0.92-2.16) | 1,10E-01 | 2,00E-01 |
| Network 7 | -0,1824 | SB2 ageFix | 0,83 | (0.59-1.17) | 3,00E-01 | 4,00E-01 |
| Network 8 | -0,3073 | SB2 ageFix | 0,74 | (0.49-1.10) | 1,40E-01 | 2,20E-01 |
| Network 9 | -0,1745 | SB2 ageFix | 0,84 | (0.53-1.34) | 4,60E-01 | 5,20E-01 |
| Network 11 | -0,0484 | SB2 ageFix | 0,95 | (0.74-1.23) | 7,10E-01 | 7,10E-01 |
| Network 12 | 0,1369 | SB2 ageFix | 1,15 | (0.90-1.46) | 2,70E-01 | 3,80E-01 |
| Network 16 | -0,1389 | SB2 ageFix | 0,87 | (0.70-1.08) | 2,10E-01 | 3,20E-01 |
| Network 19 | -0,6633 | SB2 ageFix | 0,52 | (0.37-0.72) | 8,90E-05 | 5,10E-04 |
| Network 20 | 0,1972 | SB2 ageFix | 1,22 | (0.77-1.93) | 4,00E-01 | 4,90E-01 |
| Network 24 | -0,0801 | SB2 ageFix | 0,92 | (0.67-1.28) | 6,30E-01 | 6,60E-01 |
| Network 25 | 0,0950 | SB2 ageFix | 1,10 | (0.85-1.42) | 4,70E-01 | 5,20E-01 |
| Module Age + Sex | 0,40778042 | DFH Final | 1,50 | | | |
| Module Metamix | 0,576792213 | DFH Final | 1,78 | | | |
| Module alpha diversity | 0 | DFH Final | -* | | | |
| Module beta diversity | 0 | DFH Final | -* | | | |
| Module relative abundance | 0 | DFH Final | -* | | | |
| Module curated1 | 0,000423023 | DFH Final | -* | | | |
| Module curated2 | 0 | DFH Final | -* | | | |
| Module trinary family + phylum | 0 | DFH Final | -* | | | |

*Could not be intepreted

**Supplementary Table 5. The taxonomic level profile of each co-abundance network**

| Domain | Phylum | Class | Order | Family | Genus | Species | Network |
|---|---|---|---|---|---|---|---|
| *k__Bacteria* | *p__Actinobacteria* | *c__Actinobacteria* | *o__Actinomycetales* | *f__Actinomycetaceae* | *g__Actinomyces* | *s__Actinomyces_graevenitzii* | *1* |
| *k__Bacteria* | *p__Actinobacteria* | *c__Actinobacteria* | *o__Bifidobacteriales* | *f__Bifidobacteriaceae* | *g__Bifidobacterium* | *s__Bifidobacterium_magnum* | *1* |
| *k__Bacteria* | *p__Actinobacteria* | *c__Coriobacteriia* | *o__Coriobacteriales* | *f__Atopobiaceae* | *g__Olsenella* | *s__Olsenella_profusa* | *1* |
| *k__Bacteria* | *p__Actinobacteria* | *c__Coriobacteriia* | *o__Coriobacteriales* | *f__Atopobiaceae* | *g__Olsenella* | *s__Olsenella_scatoligenes* | *1* |
| *k__Bacteria* | *p__Actinobacteria* | *c__Coriobacteriia* | *o__Coriobacteriales* | *f__Atopobiaceae* | *g__Olsenella* | *s__Olsenella_sp._SIT9* | *1* |
| *k__Bacteria* | *p__Actinobacteria* | *c__Coriobacteriia* | *o__Coriobacteriales* | *f__Atopobiaceae* | *g__Olsenella* | *s__Olsenella_sp._oral_taxon_809* | *1* |
| *k__Bacteria* | *p__Actinobacteria* | *c__Coriobacteriia* | *o__Coriobacteriales* | *f__Atopobiaceae* | *g__Olsenella* | *s__Olsenella_uli* | *1* |
| *k__Bacteria* | *p__Actinobacteria* | *c__Coriobacteriia* | *o__Coriobacteriales* | *f__Coriobacteriaceae* | *g__Collinsella* | *s__Collinsella_aerofaciens* | *1* |
| *k__Bacteria* | *p__Actinobacteria* | *c__Coriobacteriia* | *o__Coriobacteriales* | *f__Coriobacteriaceae* | *g__Collinsella* | *s__Collinsella_intestinalis* | *1* |
| *k__Bacteria* | *p__Actinobacteria* | *c__Coriobacteriia* | *o__Coriobacteriales* | *f__Coriobacteriaceae* | *g__Collinsella* | *s__Collinsella_sp._MS5* | *1* |
| *k__Bacteria* | *p__Actinobacteria* | *c__Coriobacteriia* | *o__Coriobacteriales* | *f__Coriobacteriaceae* | *g__Collinsella* | *s__Collinsella_stercoris* | *1* |
| *k__Bacteria* | *p__Actinobacteria* | *c__Coriobacteriia* | *o__Coriobacteriales* | *f__Coriobacteriaceae* | *g__Collinsella* | *s__Collinsella_tanakaei* | *1* |
| *k__Bacteria* | *p__Actinobacteria* | *c__Coriobacteriia* | *o__Coriobacteriales* | *f__Coriobacteriaceae* | *g__Enorma* | *s__Enorma_massiliensis* | *1* |
| *k__Bacteria* | *p__Actinobacteria* | *c__Coriobacteriia* | *o__Coriobacteriales* | *f__Coriobacteriaceae* | *g__Enorma* | *s__Enorma_timonensis* | *1* |
| *k__Bacteria* | *p__Actinobacteria* | *c__Coriobacteriia* | *o__Coriobacteriales* | *f__Coriobacteriaceae* | *g__Senegalimassilia* | *s__Senegalimassilia_anaerobia* | *1* |
| *k__Bacteria* | *p__Actinobacteria* | *c__Coriobacteriia* | *o__Eggerthellales* | *f__Eggerthellaceae* | *g__Slackia* | *s__Slackia_heliotrinireducens* | *1* |
| *k__Bacteria* | *p__Actinobacteria* | *c__Coriobacteriia* | *o__Eggerthellales* | *f__Eggerthellaceae* | *g__Slackia* | *s__Slackia_piriformis* | *1* |
| *k__Bacteria* | *p__Actinobacteria* | *c__Actinobacteria* | *o__Actinomycetales* | *f__Actinomycetaceae* | *g__Actinomyces* | *s__Actinomyces_odontolyticus* | *2* |
| *k__Bacteria* | *p__Actinobacteria* | *c__Actinobacteria* | *o__Actinomycetales* | *f__Actinomycetaceae* | *g__Actinomyces* | *s__Actinomyces_oris* | *2* |
| *k__Bacteria* | *p__Actinobacteria* | *c__Actinobacteria* | *o__Actinomycetales* | *f__Actinomycetaceae* | *g__Trueperella* | *s__Trueperella_pyogenes* | *2* |
| *k__Bacteria* | *p__Actinobacteria* | *c__Actinobacteria* | *o__Bifidobacteriales* | *f__Bifidobacteriaceae* | *g__Bifidobacterium* | *s__Bifidobacterium_adolescentis* | *3* |
| *k__Bacteria* | *p__Actinobacteria* | *c__Actinobacteria* | *o__Bifidobacteriales* | *f__Bifidobacteriaceae* | *g__Bifidobacterium* | *s__Bifidobacterium_aesculapii* | *3* |
| *k__Bacteria* | *p__Actinobacteria* | *c__Actinobacteria* | *o__Bifidobacteriales* | *f__Bifidobacteriaceae* | *g__Bifidobacterium* | *s__Bifidobacterium_angulatum* | *3* |
| *k__Bacteria* | *p__Actinobacteria* | *c__Actinobacteria* | *o__Bifidobacteriales* | *f__Bifidobacteriaceae* | *g__Bifidobacterium* | *s__Bifidobacterium_biavatii* | *3* |
| *k__Bacteria* | *p__Actinobacteria* | *c__Actinobacteria* | *o__Bifidobacteriales* | *f__Bifidobacteriaceae* | *g__Bifidobacterium* | *s__Bifidobacterium_bifidum* | *3* |
| *k__Bacteria* | *p__Actinobacteria* | *c__Actinobacteria* | *o__Bifidobacteriales* | *f__Bifidobacteriaceae* | *g__Bifidobacterium* | *s__Bifidobacterium_breve* | *3* |
| *k__Bacteria* | *p__Actinobacteria* | *c__Actinobacteria* | *o__Bifidobacteriales* | *f__Bifidobacteriaceae* | *g__Bifidobacterium* | *s__Bifidobacterium_callitrichos* | *3* |
| *k__Bacteria* | *p__Actinobacteria* | *c__Actinobacteria* | *o__Bifidobacteriales* | *f__Bifidobacteriaceae* | *g__Bifidobacterium* | *s__Bifidobacterium_dentium* | *3* |
| *k__Bacteria* | *p__Actinobacteria* | *c__Actinobacteria* | *o__Bifidobacteriales* | *f__Bifidobacteriaceae* | *g__Bifidobacterium* | *s__Bifidobacterium_gallinarum* | *3* |
| *k__Bacteria* | *p__Actinobacteria* | *c__Actinobacteria* | *o__Bifidobacteriales* | *f__Bifidobacteriaceae* | *g__Bifidobacterium* | *s__Bifidobacterium_longum* | *3* |
| *k__Bacteria* | *p__Actinobacteria* | *c__Actinobacteria* | *o__Bifidobacteriales* | *f__Bifidobacteriaceae* | *g__Bifidobacterium* | *s__Bifidobacterium_reuteri* | *3* |
| *k__Bacteria* | *p__Actinobacteria* | *c__Actinobacteria* | *o__Bifidobacteriales* | *f__Bifidobacteriaceae* | *g__Bifidobacterium* | *s__Bifidobacterium_saguini* | *3* |
| *k__Bacteria* | *p__Actinobacteria* | *c__Actinobacteria* | *o__Bifidobacteriales* | *f__Bifidobacteriaceae* | *g__Bifidobacterium* | *s__Bifidobacterium_scardovii* | *3* |
| *k__Bacteria* | *p__Actinobacteria* | *c__Actinobacteria* | *o__Bifidobacteriales* | *f__Bifidobacteriaceae* | *g__Bifidobacterium* | *s__Bifidobacterium_stellenboschense* | *3* |
| *k__Bacteria* | *p__Actinobacteria* | *c__Actinobacteria* | *o__Micrococcales* | *f__Cellulomonadacea* | *g__Cellulomonas* | *s__Cellulomonas_carbonis* | *4* |
| *k__Bacteria* | *p__Bacteroidetes* | *c__Bacteroidia* | *o__Bacteroidales* | *f__Porphyromonadace* | *g__Porphyromonas* | *s__Porphyromonas_somerae* | *4* |
| *k__Bacteria* | *p__Bacteroidetes* | *c__Bacteroidia* | *o__Bacteroidales* | *f__Prevotellaceae* | *g__Prevotella* | *s__Prevotella_baroniae* | *4* |
| *k__Bacteria* | *p__Bacteroidetes* | *c__Bacteroidia* | *o__Bacteroidales* | *f__Prevotellaceae* | *g__Prevotella* | *s__Prevotella_bergensis* | *4* |
| *k__Bacteria* | *p__Bacteroidetes* | *c__Bacteroidia* | *o__Bacteroidales* | *f__Prevotellaceae* | *g__Prevotella* | *s__Prevotella_bivia* | *4* |
| *k__Bacteria* | *p__Bacteroidetes* | *c__Bacteroidia* | *o__Bacteroidales* | *f__Prevotellaceae* | *g__Prevotella* | *s__Prevotella_brevis* | *4* |
| *k__Bacteria* | *p__Bacteroidetes* | *c__Bacteroidia* | *o__Bacteroidales* | *f__Prevotellaceae* | *g__Prevotella* | *s__Prevotella_buccae* | *4* |
| *k__Bacteria* | *p__Bacteroidetes* | *c__Bacteroidia* | *o__Bacteroidales* | *f__Prevotellaceae* | *g__Prevotella* | *s__Prevotella_copri* | *4* |
| *k__Bacteria* | *p__Bacteroidetes* | *c__Bacteroidia* | *o__Bacteroidales* | *f__Prevotellaceae* | *g__Prevotella* | *s__Prevotella_corporis* | *4* |
| *k__Bacteria* | *p__Bacteroidetes* | *c__Bacteroidia* | *o__Bacteroidales* | *f__Prevotellaceae* | *g__Prevotella* | *s__Prevotella_dentalis* | *4* |
| *k__Bacteria* | *p__Bacteroidetes* | *c__Bacteroidia* | *o__Bacteroidales* | *f__Prevotellaceae* | *g__Prevotella* | *s__Prevotella_dentasini* | *4* |
| *k__Bacteria* | *p__Bacteroidetes* | *c__Bacteroidia* | *o__Bacteroidales* | *f__Prevotellaceae* | *g__Prevotella* | *s__Prevotella_denticola* | *4* |
| *k__Bacteria* | *p__Bacteroidetes* | *c__Bacteroidia* | *o__Bacteroidales* | *f__Prevotellaceae* | *g__Prevotella* | *s__Prevotella_fusca* | *4* |
| *k__Bacteria* | *p__Bacteroidetes* | *c__Bacteroidia* | *o__Bacteroidales* | *f__Prevotellaceae* | *g__Prevotella* | *s__Prevotella_multisaccharivorax* | *4* |
| *k__Bacteria* | *p__Bacteroidetes* | *c__Bacteroidia* | *o__Bacteroidales* | *f__Prevotellaceae* | *g__Prevotella* | *s__Prevotella_sp._AGR2160* | *4* |
| *k__Bacteria* | *p__Bacteroidetes* | *c__Bacteroidia* | *o__Bacteroidales* | *f__Prevotellaceae* | *g__Prevotella* | *s__Prevotella_sp._P4-76* | *4* |
| *k__Bacteria* | *p__Bacteroidetes* | *c__Bacteroidia* | *o__Bacteroidales* | *f__Prevotellaceae* | *g__Prevotella* | *s__Prevotella_stercorea* | *4* |
| *k__Bacteria* | *p__Actinobacteria* | *c__Coriobacteriia* | *o__Eggerthellales* | *f__Eggerthellaceae* | *g__Adlercreutzia* | *s__Adlercreutzia_equolifaciens* | *5* |
| *k__Bacteria* | *p__Actinobacteria* | *c__Coriobacteriia* | *o__Eggerthellales* | *f__Eggerthellaceae* | *g__Eggerthella* | *s__Eggerthella_sp._YY7918* | *5* |
| *k__Bacteria* | *p__Actinobacteria* | *c__Coriobacteriia* | *o__Eggerthellales* | *f__Eggerthellaceae* | *g__Enterorhabdus* | *s__Enterorhabdus_caecimuris* | *5* |
| *k__Bacteria* | *p__Actinobacteria* | *c__Coriobacteriia* | *o__Eggerthellales* | *f__Eggerthellaceae* | *g__Enterorhabdus* | *s__Enterorhabdus_mucosicola* | *5* |
| *k__Bacteria* | *p__Actinobacteria* | *c__Coriobacteriia* | *o__Eggerthellales* | *f__Eggerthellaceae* | *g__Eggerthella* | *s__Eggerthella_lenta* | *6* |
| *k__Bacteria* | *p__Actinobacteria* | *c__Coriobacteriia* | *o__Eggerthellales* | *f__Eggerthellaceae* | *g__Gordonibacter* | *s__Gordonibacter_pamelaeae* | *6* |
| *k__Bacteria* | *p__Firmicutes* | *c__Clostridia* | *o__Clostridiales* | *f__Clostridiaceae* | *g__Clostridium* | *s__Clostridium_sp._KLE_1755* | *6* |
| *k__Bacteria* | *p__Firmicutes* | *c__Clostridia* | *o__Clostridiales* | *f__Clostridiaceae* | *g__Hungatella* | *s__Hungatella_hathewayi* | *6* |
| *k__Bacteria* | *p__Firmicutes* | *c__Clostridia* | *o__Clostridiales* | *f__Lachnospiraceae* | *g__Anaerostipes* | *s__Anaerostipes_caccae* | *6* |
| *k__Bacteria* | *p__Firmicutes* | *c__Clostridia* | *o__Clostridiales* | *f__Lachnospiraceae* | *g__Eisenbergiella* | *s__Eisenbergiella_tayi* | *6* |
| *k__Bacteria* | *p__Firmicutes* | *c__Clostridia* | *o__Clostridiales* | *f__Lachnospiraceae* | *g__Lachnoclostridium* | *s__[Clostridium]_asparagiforme* | *6* |
| *k__Bacteria* | *p__Firmicutes* | *c__Clostridia* | *o__Clostridiales* | *f__Lachnospiraceae* | *g__Lachnoclostridium* | *s__[Clostridium]_bolteae* | *6* |
| *k__Bacteria* | *p__Firmicutes* | *c__Clostridia* | *o__Clostridiales* | *f__Lachnospiraceae* | *g__Lachnoclostridium* | *s__[Clostridium]_citroniae* | *6* |
| *k__Bacteria* | *p__Firmicutes* | *c__Clostridia* | *o__Clostridiales* | *f__Lachnospiraceae* | *g__Lachnoclostridium* | *s__[Clostridium]_symbiosum* | *6* |
| *k__Bacteria* | *p__Firmicutes* | *c__Clostridia* | *o__Clostridiales* | *f__Lachnospiraceae* | *g__Sellimonas* | *s__Sellimonas_intestinalis* | *6* |
| *k__Bacteria* | *p__Firmicutes* | *c__Clostridia* | *o__Clostridiales* | *f__Ruminococcaceae* | *g__Ruthenibacterium* | *s__Ruthenibacterium_lactatiformans* | *6* |
| *k__Bacteria* | *p__Firmicutes* | *c__Erysipelotrichia* | *o__Erysipelotrichales* | *f__Erysipelotrichacea* | *g__Dielma* | *s__Dielma_fastidiosa* | *6* |
| *k__Bacteria* | *p__Firmicutes* | *c__Erysipelotrichia* | *o__Erysipelotrichales* | *f__Erysipelotrichacea* | *g__Erysipelatoclostridium* | *s__Erysipelatoclostridium_ramosum* | *6* |
| *k__Bacteria* | *p__Firmicutes* | *c__Erysipelotrichia* | *o__Erysipelotrichales* | *f__Erysipelotrichacea* | *g__Erysipelatoclostridium* | *s__[Clostridium]_innocuum* | *6* |
| *k__Bacteria* | *p__Firmicutes* | *c__Erysipelotrichia* | *o__Erysipelotrichales* | *f__Erysipelotrichacea* | *g__Erysipelatoclostridium* | *s__[Clostridium]_spiroforme* | *6* |
| *k__Bacteria* | *p__Firmicutes* | *c__Erysipelotrichia* | *o__Erysipelotrichales* | *f__Erysipelotrichacea* | *g__Holdemania* | *s__Holdemania_massiliensis* | *6* |
| *k__Bacteria* | *p__Bacteroidetes* | *c__Bacteroidia* | *o__Bacteroidales* | *f__Bacteroidaceae* | *g__Bacteroides* | *s__Bacteroides_barnesiae* | *7* |
| *k__Bacteria* | *p__Bacteroidetes* | *c__Bacteroidia* | *o__Bacteroidales* | *f__Bacteroidaceae* | *g__Bacteroides* | *s__Bacteroides_coprocola* | *7* |
| *k__Bacteria* | *p__Bacteroidetes* | *c__Bacteroidia* | *o__Bacteroidales* | *f__Bacteroidaceae* | *g__Bacteroides* | *s__Bacteroides_coprophilus* | *7* |
| *k__Bacteria* | *p__Bacteroidetes* | *c__Bacteroidia* | *o__Bacteroidales* | *f__Bacteroidaceae* | *g__Bacteroides* | *s__Bacteroides_plebeius* | *7* |
| *k__Bacteria* | *p__Bacteroidetes* | *c__Bacteroidia* | *o__Bacteroidales* | *f__Bacteroidaceae* | *g__Bacteroides* | *s__Bacteroides_salanitronis* | *7* |
| *k__Bacteria* | *p__Bacteroidetes* | *c__Bacteroidia* | *o__Bacteroidales* | *f__Barnesiellaceae* | *g__Barnesiella* | *s__Barnesiella_viscericola* | *7* |
| *k__Bacteria* | *p__Bacteroidetes* | *c__Bacteroidia* | *o__Bacteroidales* | *f__Barnesiellaceae* | *g__Coprobacter* | *s__Coprobacter_secundus* | *7* |
| *k__Bacteria* | *p__Bacteroidetes* | *c__Bacteroidia* | *o__Bacteroidales* | *f__Prevotellaceae* | *g__Prevotella* | *s__Prevotella_sp._109* | *7* |
| *k__Bacteria* | *p__Bacteroidetes* | *c__Bacteroidia* | *o__Bacteroidales* | *f__Rikenellaceae* | *g__Rikenella* | *s__Rikenella_microfusus* | *7* |
| *k__Bacteria* | *p__Bacteroidetes* | *c__Bacteroidia* | *o__Bacteroidales* | *f__Bacteroidaceae* | *g__Bacteroides* | *s__Bacteroides_cellulosilyticus* | *8* |
| *k__Bacteria* | *p__Bacteroidetes* | *c__Bacteroidia* | *o__Bacteroidales* | *f__Bacteroidaceae* | *g__Bacteroides* | *s__Bacteroides_coprosuis* | *8* |
| *k__Bacteria* | *p__Bacteroidetes* | *c__Bacteroidia* | *o__Bacteroidales* | *f__Bacteroidaceae* | *g__Bacteroides* | *s__Bacteroides_faecichinchillae* | *8* |
| *k__Bacteria* | *p__Bacteroidetes* | *c__Bacteroidia* | *o__Bacteroidales* | *f__Bacteroidaceae* | *g__Bacteroides* | *s__Bacteroides_fluxus* | *8* |
| *k__Bacteria* | *p__Bacteroidetes* | *c__Bacteroidia* | *o__Bacteroidales* | *f__Bacteroidaceae* | *g__Bacteroides* | *s__Bacteroides_fragilis* | *8* |
| *k__Bacteria* | *p__Bacteroidetes* | *c__Bacteroidia* | *o__Bacteroidales* | *f__Bacteroidaceae* | *g__Bacteroides* | *s__Bacteroides_graminisolvens* | *8* |
| *k__Bacteria* | *p__Bacteroidetes* | *c__Bacteroidia* | *o__Bacteroidales* | *f__Bacteroidaceae* | *g__Bacteroides* | *s__Bacteroides_neonati* | *8* |
| *k__Bacteria* | *p__Bacteroidetes* | *c__Bacteroidia* | *o__Bacteroidales* | *f__Bacteroidaceae* | *g__Bacteroides* | *s__Bacteroides_oleiciplenus* | *8* |
| *k__Bacteria* | *p__Bacteroidetes* | *c__Bacteroidia* | *o__Bacteroidales* | *f__Bacteroidaceae* | *g__Bacteroides* | *s__Bacteroides_ovatus* | *8* |
| *k__Bacteria* | *p__Bacteroidetes* | *c__Bacteroidia* | *o__Bacteroidales* | *f__Bacteroidaceae* | *g__Bacteroides* | *s__Bacteroides_paurosaccharolyticus* | *8* |
| *k__Bacteria* | *p__Bacteroidetes* | *c__Bacteroidia* | *o__Bacteroidales* | *f__Bacteroidaceae* | *g__Bacteroides* | *s__Bacteroides_pyogenes* | *8* |
| *k__Bacteria* | *p__Bacteroidetes* | *c__Bacteroidia* | *o__Bacteroidales* | *f__Bacteroidaceae* | *g__Bacteroides* | *s__Bacteroides_reticulotermitis* | *8* |
| *k__Bacteria* | *p__Bacteroidetes* | *c__Bacteroidia* | *o__Bacteroidales* | *f__Bacteroidaceae* | *g__Bacteroides* | *s__Bacteroides_salyersiae* | *8* |
| *k__Bacteria* | *p__Bacteroidetes* | *c__Bacteroidia* | *o__Bacteroidales* | *f__Bacteroidaceae* | *g__Bacteroides* | *s__Bacteroides_stercoris* | *8* |
| *k__Bacteria* | *p__Bacteroidetes* | *c__Bacteroidia* | *o__Bacteroidales* | *f__Bacteroidaceae* | *g__Bacteroides* | *s__Bacteroides_thetaiotaomicron* | *8* |
| *k__Bacteria* | *p__Bacteroidetes* | *c__Bacteroidia* | *o__Bacteroidales* | *f__Bacteroidaceae* | *g__Bacteroides* | *s__Bacteroides_uniformis* | *8* |
| *k__Bacteria* | *p__Bacteroidetes* | *c__Bacteroidia* | *o__Bacteroidales* | *f__Bacteroidaceae* | *g__Bacteroides* | *s__Bacteroides_vulgatus* | *8* |
| *k__Bacteria* | *p__Bacteroidetes* | *c__Bacteroidia* | *o__Bacteroidales* | *f__Barnesiellaceae* | *g__Coprobacter* | *s__Coprobacter_fastidiosus* | *8* |
| *k__Bacteria* | *p__Bacteroidetes* | *c__Bacteroidia* | *o__Bacteroidales* | *f__Odoribacteraceae* | *g__Odoribacter* | *s__Odoribacter_laneus* | *8* |
| *k__Bacteria* | *p__Bacteroidetes* | *c__Bacteroidia* | *o__Bacteroidales* | *f__Porphyromonadace* | *g__Sanguibacteroides* | *s__Sanguibacteroides_justesenii* | *8* |
| *k__Bacteria* | *p__Bacteroidetes* | *c__Bacteroidia* | *o__Bacteroidales* | *f__Tannerellaceae* | *g__Parabacteroides* | *s__Parabacteroides_distasonis* | *8* |
| *k__Bacteria* | *p__Bacteroidetes* | *c__Bacteroidia* | *o__Bacteroidales* | *f__Tannerellaceae* | *g__Parabacteroides* | *s__Parabacteroides_johnsonii* | *8* |
| *k__Bacteria* | *p__Bacteroidetes* | *c__Bacteroidia* | *o__Bacteroidales* | *f__Barnesiellaceae* | *g__Barnesiella* | *s__Barnesiella_intestinihominis* | *9* |
| *k__Bacteria* | *p__Bacteroidetes* | *c__Bacteroidia* | *o__Bacteroidales* | *f__Odoribacteraceae* | *g__Butyricimonas* | *s__Butyricimonas_synergistica* | *9* |
| *k__Bacteria* | *p__Bacteroidetes* | *c__Bacteroidia* | *o__Bacteroidales* | *f__Odoribacteraceae* | *g__Butyricimonas* | *s__Butyricimonas_virosa* | *9* |
| *k__Bacteria* | *p__Bacteroidetes* | *c__Bacteroidia* | *o__Bacteroidales* | *f__Odoribacteraceae* | *g__Odoribacter* | *s__Odoribacter_splanchnicus* | *9* |
| *k__Bacteria* | *p__Bacteroidetes* | *c__Bacteroidia* | *o__Bacteroidales* | *f__Prevotellaceae* | *g__Paraprevotella* | *s__Paraprevotella_xylaniphila* | *9* |
| *k__Bacteria* | *p__Firmicutes* | *c__Clostridia* | *o__Clostridiales* | *f__Clostridiaceae* | *g__Butyricicoccus* | *s__Butyricicoccus_desmolans* | *9* |
| *k__Bacteria* | *p__Firmicutes* | *c__Clostridia* | *o__Clostridiales* | *f__Lachnospiraceae* | *g__Blautia* | *s__Blautia_hydrogenotrophica* | *9* |
| *k__Bacteria* | *p__Firmicutes* | *c__Clostridia* | *o__Clostridiales* | *f__Lachnospiraceae* | *g__Blautia* | *s__Blautia_schinkii* | *9* |
| *k__Bacteria* | *p__Firmicutes* | *c__Clostridia* | *o__Clostridiales* | *f__Lachnospiraceae* | *g__Faecalicatena* | *s__Faecalicatena_contorta* | *9* |
| *k__Bacteria* | *p__Firmicutes* | *c__Clostridia* | *o__Clostridiales* | *f__Lachnospiraceae* | *g__Marvinbryantia* | *s__Marvinbryantia_formatexigens* | *9* |
| *k__Bacteria* | *p__Firmicutes* | *c__Erysipelotrichia* | *o__Erysipelotrichales* | *f__Erysipelotrichacea* | *g__Faecalitalea* | *s__Faecalitalea_cylindroides* | *9* |
| *k__Bacteria* | *p__Bacteroidetes* | *c__Bacteroidia* | *o__Bacteroidales* | *f__Porphyromonadace* | *g__Porphyromonas* | *s__Porphyromonas_crevioricanis* | *10* |
| *k__Bacteria* | *p__Bacteroidetes* | *c__Bacteroidia* | *o__Bacteroidales* | *f__Prevotellaceae* | *g__Alloprevotella* | *s__Alloprevotella_tannerae* | *10* |
| *k__Bacteria* | *p__Bacteroidetes* | *c__Bacteroidia* | *o__Bacteroidales* | *f__Prevotellaceae* | *g__Prevotella* | *s__Prevotella_amnii* | *10* |
| *k__Bacteria* | *p__Bacteroidetes* | *c__Bacteroidia* | *o__Bacteroidales* | *f__Prevotellaceae* | *g__Prevotella* | *s__Prevotella_buccalis* | *10* |
| *k__Bacteria* | *p__Bacteroidetes* | *c__Bacteroidia* | *o__Bacteroidales* | *f__Prevotellaceae* | *g__Prevotella* | *s__Prevotella_disiens* | *10* |
| *k__Bacteria* | *p__Bacteroidetes* | *c__Bacteroidia* | *o__Bacteroidales* | *f__Prevotellaceae* | *g__Prevotella* | *s__Prevotella_multiformis* | *10* |
| *k__Bacteria* | *p__Bacteroidetes* | *c__Bacteroidia* | *o__Bacteroidales* | *f__Prevotellaceae* | *g__Prevotella* | *s__Prevotella_sp._10(H)* | *10* |
| *k__Bacteria* | *p__Bacteroidetes* | *c__Bacteroidia* | *o__Bacteroidales* | *f__Prevotellaceae* | *g__Prevotella* | *s__Prevotella_sp._HUN102* | *10* |
| *k__Bacteria* | *p__Bacteroidetes* | *c__Bacteroidia* | *o__Bacteroidales* | *f__Prevotellaceae* | *g__Prevotella* | *s__Prevotella_sp._S7_MS_2* | *10* |
| *k__Bacteria* | *p__Bacteroidetes* | *c__Bacteroidia* | *o__Bacteroidales* | *f__Prevotellaceae* | *g__Prevotella* | *s__Prevotella_timonensis* | *10* |
| *k__Bacteria* | *p__Bacteroidetes* | *c__Bacteroidia* | *o__Bacteroidales* | *f__Tannerellaceae* | *g__Parabacteroides* | *s__Parabacteroides_goldsteinii* | *10* |
| *k__Bacteria* | *p__Bacteroidetes* | *c__Bacteroidia* | *o__Bacteroidales* | *f__Tannerellaceae* | *g__Tannerella* | *s__Tannerella_forsythia* | *10* |
| *k__Bacteria* | *p__Bacteroidetes* | *c__Bacteroidia* | *o__Bacteroidales* | *f__Rikenellaceae* | *g__Alistipes* | *s__Alistipes_finegoldii* | *11* |
| *k__Bacteria* | *p__Bacteroidetes* | *c__Bacteroidia* | *o__Bacteroidales* | *f__Rikenellaceae* | *g__Alistipes* | *s__Alistipes_indistinctus* | *11* |

| | | | | | | | |
|---|---|---|---|---|---|---|---|
| k__Bacteria | p__Bacteroidetes | c__Bacteroidia | o__Bacteroidales | f__Rikenellaceae | g__Alistipes | s__Alistipes_obesi | 11 |
| k__Bacteria | p__Bacteroidetes | c__Bacteroidia | o__Bacteroidales | f__Rikenellaceae | g__Alistipes | s__Alistipes_putredinis | 11 |
| k__Bacteria | p__Bacteroidetes | c__Bacteroidia | o__Bacteroidales | f__Rikenellaceae | g__Alistipes | s__Alistipes_senegalensis | 11 |
| k__Bacteria | p__Bacteroidetes | c__Bacteroidia | o__Bacteroidales | f__Rikenellaceae | g__Alistipes | s__Alistipes_shahii | 11 |
| k__Bacteria | p__Bacteroidetes | c__Bacteroidia | o__Bacteroidales | f__Rikenellaceae | g__Alistipes | s__Alistipes_timonensis | 11 |
| k__Bacteria | p__Firmicutes | c__Bacilli | o__Lactobacillales | f__Lactobacillaceae | g__Sharpea | s__Sharpea_azabuensis | 12 |
| k__Bacteria | p__Firmicutes | c__Erysipelotrichia | o__Erysipelotrichales | f__Erysipelotrichacea | g__Catenibacterium | s__Catenibacterium_mitsuokai | 12 |
| k__Bacteria | p__Firmicutes | c__Bacilli | o__Lactobacillales | f__Streptococcaceae | g__Streptococcus | s__Streptococcus_mitis | 13 |
| k__Bacteria | p__Firmicutes | c__Bacilli | o__Lactobacillales | f__Streptococcaceae | g__Streptococcus | s__Streptococcus_parasanguinis | 13 |
| k__Bacteria | p__Firmicutes | c__Bacilli | o__Lactobacillales | f__Streptococcaceae | g__Streptococcus | s__Streptococcus_salivarius | 13 |
| k__Bacteria | p__Firmicutes | c__Bacilli | o__Lactobacillales | f__Streptococcaceae | g__Streptococcus | s__Streptococcus_thermophilus | 13 |
| k__Bacteria | p__Firmicutes | c__Negativicutes | o__Veillonellales | f__Veillonellaceae | g__Veillonella | s__Veillonella_parvula | 13 |
| k__Bacteria | p__Firmicutes | c__Clostridia | o__Clostridiales | f__Catabacteriaceae | g__Catabacter | s__Catabacter_hongkongensis | 14 |
| k__Bacteria | p__Firmicutes | c__Clostridia | o__Clostridiales | f__Heliobacteriaceae | g__Heliobacterium | s__Heliobacterium_modesticaldum | 14 |
| k__Bacteria | p__Firmicutes | c__Clostridia | o__Clostridiales | f__Lachnospiraceae | g__Lachnoanaerobaculum | s__Lachnoanaerobaculum_sp._OBRC5-5 | 14 |
| k__Bacteria | p__Firmicutes | c__Clostridia | o__Clostridiales | f__Oscillospiraceae | g__Oscillibacter | s__Oscillibacter_ruminantium | 14 |
| k__Bacteria | p__Firmicutes | c__Clostridia | o__Clostridiales | f__Oscillospiraceae | g__Oscillibacter | s__Oscillibacter_sp._1-3 | 14 |
| k__Bacteria | p__Firmicutes | c__Clostridia | o__Clostridiales | f__Oscillospiraceae | g__Oscillibacter | s__Oscillibacter_sp._ER4 | 14 |
| k__Bacteria | p__Firmicutes | c__Clostridia | o__Clostridiales | f__Oscillospiraceae | g__Oscillibacter | s__Oscillibacter_valericigenes | 14 |
| k__Bacteria | p__Firmicutes | c__Clostridia | o__Clostridiales | f__Ruminococcaceae | g__Candidatus_Soleaferrea | s__Candidatus_Soleaferrea_massiliensis | 14 |
| k__Bacteria | p__Firmicutes | c__Clostridia | o__Clostridiales | f__Ruminococcaceae | g__Ethanoligenens | s__Ethanoligenens_harbinense | 14 |
| k__Bacteria | p__Firmicutes | c__Clostridia | o__Clostridiales | f__Ruminococcaceae | g__Mageeibacillus | s__Mageeibacillus_indolicus | 14 |
| k__Bacteria | p__Firmicutes | c__Clostridia | o__Clostridiales | f__Ruminococcaceae | g__Ruminiclostridium | s__[Clostridium]_cellulosi | 14 |
| k__Bacteria | p__Firmicutes | c__Clostridia | o__Clostridiales | f__Ruminococcaceae | g__Ruminiclostridium | s__[Clostridium]_sporosphaeroides | 14 |
| k__Bacteria | p__Firmicutes | c__Clostridia | o__Clostridiales | f__Ruminococcaceae | g__Ruminiclostridium | s__[Clostridium]_viride | 14 |
| k__Bacteria | p__Firmicutes | c__Clostridia | o__Clostridiales | f__Ruminococcaceae | g__Ruminococcus | s__Ruminococcus_champanellensis | 14 |
| k__Bacteria | p__Firmicutes | c__Clostridia | o__Thermoanaerobacterales | f__Thermoanaerobact | g__Mahella | s__Mahella_australiensis | 14 |
| k__Bacteria | p__Firmicutes | c__Negativicutes | o__Acidaminococcales | f__Acidaminococcace | g__Acidaminococcus | s__Acidaminococcus_fermentans | 14 |
| k__Bacteria | p__Firmicutes | c__Negativicutes | o__Selenomonadales | f__Selenomonadacea | g__Mitsuokella | s__Mitsuokella_multacida | 14 |
| k__Bacteria | p__Firmicutes | c__Negativicutes | o__Selenomonadales | f__Selenomonadacea | g__Selenomonas | s__Selenomonas_bovis | 14 |
| k__Bacteria | p__Firmicutes | c__Negativicutes | o__Selenomonadales | f__Selenomonadacea | g__Selenomonas | s__Selenomonas_ruminantium | 14 |
| k__Bacteria | p__Firmicutes | c__Tissierellia | o__Tissierellales | f__Peptoniphilaceae | g__Peptoniphilus | s__Peptoniphilus_sp._BV3C26 | 14 |
| k__Bacteria | p__Proteobacteria | c__Deltaproteobacteria | o__Desulfovibrionales | f__Desulfovibrionace | g__Desulfovibrio | s__Desulfovibrio_desulfuricans | 14 |
| k__Bacteria | p__Synergistetes | c__Synergistia | o__Synergistales | f__Synergistaceae | g__Pyramidobacter | s__Pyramidobacter_piscolens | 14 |
| k__Bacteria | p__Firmicutes | c__Clostridia | o__Clostridiales | f__Clostridiaceae | g__Butyricicoccus | s__Butyricicoccus_pullicaecorum | 15 |
| k__Bacteria | p__Firmicutes | c__Clostridia | o__Clostridiales | f__Clostridiaceae | g__Clostridium | s__Clostridium_phoceensis | 15 |
| k__Bacteria | p__Firmicutes | c__Clostridia | o__Clostridiales | f__Clostridiaceae | g__Clostridium | s__Clostridium_sp._M62/1 | 15 |
| k__Bacteria | p__Firmicutes | c__Clostridia | o__Clostridiales | f__Eubacteriaceae | g__Eubacterium | s__Eubacterium_limosum | 15 |
| k__Bacteria | p__Firmicutes | c__Clostridia | o__Clostridiales | f__Eubacteriaceae | g__Eubacterium | s__Eubacterium_sp._ER2 | 15 |
| k__Bacteria | p__Firmicutes | c__Clostridia | o__Clostridiales | f__Lachnospiraceae | g__Johnsonella | s__Johnsonella_ignava | 15 |
| k__Bacteria | p__Firmicutes | c__Clostridia | o__Clostridiales | f__Lachnospiraceae | g__Tyzzerella | s__[Clostridium]_lactatifermentans | 15 |
| k__Bacteria | p__Firmicutes | c__Clostridia | o__Clostridiales | f__Oscillospiraceae | g__Oscillibacter | s__Oscillibacter_sp._KLE_1745 | 15 |
| k__Bacteria | p__Firmicutes | c__Clostridia | o__Clostridiales | f__Ruminococcaceae | g__Anaeromassilibacillus | s__Anaeromassilibacillus_senegalensis | 15 |
| k__Bacteria | p__Firmicutes | c__Clostridia | o__Clostridiales | f__Ruminococcaceae | g__Anaerotruncus | s__Anaerotruncus_colihominis | 15 |
| k__Bacteria | p__Firmicutes | c__Clostridia | o__Clostridiales | f__Ruminococcaceae | g__Anaerotruncus | s__Anaerotruncus_sp._G3(2012) | 15 |
| k__Bacteria | p__Firmicutes | c__Clostridia | o__Clostridiales | f__Ruminococcaceae | g__Bittarella | s__Bittarella_massiliensis | 15 |
| k__Bacteria | p__Firmicutes | c__Clostridia | o__Clostridiales | f__Ruminococcaceae | g__Fournierella | s__Fournierella_massiliensis | 15 |
| k__Bacteria | p__Firmicutes | c__Clostridia | o__Clostridiales | f__Ruminococcaceae | g__Phocea | s__Phocea_massiliensis | 15 |
| k__Bacteria | p__Firmicutes | c__Clostridia | o__Clostridiales | f__Ruminococcaceae | g__Ruminiclostridium | s__[Clostridium]_methylpentosum | 15 |
| k__Bacteria | p__Firmicutes | c__Clostridia | o__Clostridiales | f__Ruminococcaceae | g__Subdoligranulum | s__Subdoligranulum_variabile | 15 |
| k__Bacteria | p__Firmicutes | c__Erysipelotrichia | o__Erysipelotrichales | f__Erysipelotrichacea | g__Faecalicoccus | s__Faecalicoccus_pleomorphus | 15 |
| k__Bacteria | p__Firmicutes | c__Clostridia | o__Clostridiales | f__Clostridiaceae | g__Clostridium | s__Clostridium_disporicum | 16 |
| k__Bacteria | p__Firmicutes | c__Clostridia | o__Clostridiales | f__Peptostreptococca | g__Intestinibacter | s__Intestinibacter_bartlettii | 16 |
| k__Bacteria | p__Firmicutes | c__Clostridia | o__Clostridiales | f__Clostridiaceae | g__Clostridium | s__Clostridium_sp._KNHs205 | 17 |
| k__Bacteria | p__Firmicutes | c__Clostridia | o__Clostridiales | f__Eubacteriaceae | g__Eubacterium | s__Eubacterium_plexicaudatum | 17 |
| k__Bacteria | p__Firmicutes | c__Clostridia | o__Clostridiales | f__Eubacteriaceae | g__Eubacterium | s__[Eubacterium]_cellulosolvens | 17 |
| k__Bacteria | p__Firmicutes | c__Clostridia | o__Clostridiales | f__Lachnospiraceae | g__Faecalicatena | s__Faecalicatena_fissicatena | 17 |
| k__Bacteria | p__Firmicutes | c__Clostridia | o__Clostridiales | f__Lachnospiraceae | g__Lachnoclostridium | s__[Clostridium]_aerotolerans | 17 |
| k__Bacteria | p__Firmicutes | c__Clostridia | o__Clostridiales | f__Lachnospiraceae | g__Lachnoclostridium | s__[Clostridium]_aminophilum | 17 |
| k__Bacteria | p__Firmicutes | c__Clostridia | o__Clostridiales | f__Lachnospiraceae | g__Lachnoclostridium | s__[Clostridium]_celerecrescens | 17 |
| k__Bacteria | p__Firmicutes | c__Clostridia | o__Clostridiales | f__Lachnospiraceae | g__Lachnoclostridium | s__[Clostridium]_hylemonae | 17 |
| k__Bacteria | p__Firmicutes | c__Clostridia | o__Clostridiales | f__Lachnospiraceae | g__Robinsoniella | s__Robinsoniella_peoriensis | 17 |
| k__Bacteria | p__Firmicutes | c__Clostridia | o__Clostridiales | f__Ruminococcaceae | g__Ruminococcus | s__Ruminococcus_gauvreauii | 17 |
| k__Bacteria | p__Firmicutes | c__Clostridia | o__Clostridiales | f__Clostridiaceae | g__Clostridium | s__Clostridium_sp._KNHs209 | 18 |
| k__Bacteria | p__Firmicutes | c__Clostridia | o__Clostridiales | f__Eubacteriaceae | g__Eubacterium | s__Eubacterium_xylanophilum | 18 |
| k__Bacteria | p__Firmicutes | c__Clostridia | o__Clostridiales | f__Lachnospiraceae | g__Butyrivibrio | s__Butyrivibrio_fibrisolvens | 18 |
| k__Bacteria | p__Firmicutes | c__Clostridia | o__Clostridiales | f__Lachnospiraceae | g__Butyrivibrio | s__Butyrivibrio_hungatei | 18 |
| k__Bacteria | p__Firmicutes | c__Clostridia | o__Clostridiales | f__Lachnospiraceae | g__Butyrivibrio | s__Butyrivibrio_sp._AE3004 | 18 |
| k__Bacteria | p__Firmicutes | c__Clostridia | o__Clostridiales | f__Lachnospiraceae | g__Butyrivibrio | s__Butyrivibrio_sp._AE3006 | 18 |
| k__Bacteria | p__Firmicutes | c__Clostridia | o__Clostridiales | f__Lachnospiraceae | g__Butyrivibrio | s__Butyrivibrio_sp._MC2013 | 18 |
| k__Bacteria | p__Firmicutes | c__Clostridia | o__Clostridiales | f__Lachnospiraceae | g__Butyrivibrio | s__Butyrivibrio_sp._NC2002 | 18 |
| k__Bacteria | p__Firmicutes | c__Clostridia | o__Clostridiales | f__Lachnospiraceae | g__Butyrivibrio | s__Butyrivibrio_sp._NC3005 | 18 |
| k__Bacteria | p__Firmicutes | c__Clostridia | o__Clostridiales | f__Lachnospiraceae | g__Oribacterium | s__Oribacterium_parvum | 18 |
| k__Bacteria | p__Firmicutes | c__Clostridia | o__Clostridiales | f__Lachnospiraceae | g__Oribacterium | s__Oribacterium_sinus | 18 |
| k__Bacteria | p__Firmicutes | c__Clostridia | o__Clostridiales | f__Lachnospiraceae | g__Roseburia | s__Roseburia_hominis | 18 |
| k__Bacteria | p__Firmicutes | c__Clostridia | o__Clostridiales | f__Ruminococcaceae | g__Acetivibrio | s__Acetivibrio_ethanolgignens | 18 |
| k__Bacteria | p__Firmicutes | c__Clostridia | o__Clostridiales | f__Eubacteriaceae | g__Eubacterium | s__Eubacterium_ramulus | 19 |
| k__Bacteria | p__Firmicutes | c__Clostridia | o__Clostridiales | f__Eubacteriaceae | g__Eubacterium | s__Eubacterium_sp._AB3007 | 19 |
| k__Bacteria | p__Firmicutes | c__Clostridia | o__Clostridiales | f__Eubacteriaceae | g__Eubacterium | s__[Eubacterium]_hallii | 19 |
| k__Bacteria | p__Firmicutes | c__Clostridia | o__Clostridiales | f__Lachnospiraceae | g__Anaerostipes | s__Anaerostipes_hadrus | 19 |
| k__Bacteria | p__Firmicutes | c__Clostridia | o__Clostridiales | f__Lachnospiraceae | g__Blautia | s__Blautia_obeum | 19 |
| k__Bacteria | p__Firmicutes | c__Clostridia | o__Clostridiales | f__Lachnospiraceae | g__Blautia | s__Blautia_sp._KLE_1732 | 19 |
| k__Bacteria | p__Firmicutes | c__Clostridia | o__Clostridiales | f__Lachnospiraceae | g__Coprococcus | s__Coprococcus_comes | 19 |
| k__Bacteria | p__Firmicutes | c__Clostridia | o__Clostridiales | f__Lachnospiraceae | g__Dorea | s__Dorea_formicigenerans | 19 |
| k__Bacteria | p__Firmicutes | c__Clostridia | o__Clostridiales | f__Lachnospiraceae | g__Dorea | s__Dorea_longicatena | 19 |
| k__Bacteria | p__Firmicutes | c__Clostridia | o__Clostridiales | f__Ruminococcaceae | g__Ruminococcus | s__Ruminococcus_faecis | 19 |
| k__Bacteria | p__Firmicutes | c__Clostridia | o__Clostridiales | f__Eubacteriaceae | g__Eubacterium | s__Eubacterium_sp._14-2 | 20 |
| k__Bacteria | p__Firmicutes | c__Clostridia | o__Clostridiales | f__Eubacteriaceae | g__Eubacterium | s__Eubacterium_sp._3_1_31 | 20 |
| k__Bacteria | p__Firmicutes | c__Clostridia | o__Clostridiales | f__Lachnospiraceae | g__Blautia | s__Blautia_hansenii | 20 |
| k__Bacteria | p__Firmicutes | c__Clostridia | o__Clostridiales | f__Lachnospiraceae | g__Blautia | s__Blautia_producta | 20 |
| k__Bacteria | p__Firmicutes | c__Clostridia | o__Clostridiales | f__Lachnospiraceae | g__Blautia | s__[Ruminococcus]_gnavus | 20 |
| k__Bacteria | p__Firmicutes | c__Clostridia | o__Clostridiales | f__Lachnospiraceae | g__Blautia | s__[Ruminococcus]_torques | 20 |
| k__Bacteria | p__Firmicutes | c__Clostridia | o__Clostridiales | f__Lachnospiraceae | g__Coprococcus | s__Coprococcus_sp._HPP0048 | 20 |
| k__Bacteria | p__Firmicutes | c__Clostridia | o__Clostridiales | f__Lachnospiraceae | g__Lachnoclostridium | s__[Clostridium]_glycyrrhizinilyticum | 20 |
| k__Bacteria | p__Firmicutes | c__Clostridia | o__Clostridiales | f__Lachnospiraceae | g__Lachnoclostridium | s__[Clostridium]_scindens | 20 |
| k__Bacteria | p__Firmicutes | c__Clostridia | o__Clostridiales | f__Lachnospiraceae | g__Tyzzerella | s__Tyzzerella_nexilis | 20 |
| k__Bacteria | p__Firmicutes | c__Clostridia | o__Clostridiales | f__Peptostreptococca | g__Clostridioides | s__Clostridioides_difficile | 20 |
| k__Bacteria | p__Firmicutes | c__Clostridia | o__Clostridiales | f__Ruminococcaceae | g__Ruminococcus | s__Ruminococcus_sp._AT10 | 20 |
| k__Bacteria | p__Firmicutes | c__Erysipelotrichia | o__Erysipelotrichales | f__Erysipelotrichacea | g__Erysipelatoclostridium | s__[Clostridium]_saccharogumia | 20 |
| k__Bacteria | p__Firmicutes | c__Clostridia | o__Clostridiales | f__Eubacteriaceae | g__Eubacterium | s__[Eubacterium]_eligens | 21 |
| k__BacteriaPlasmid | p__Firmicutes | c__Clostridia | o__Clostridiales | f__Eubacteriaceae | g__Eubacterium | s__[Eubacterium]_eligens | 21 |
| k__Bacteria | p__Firmicutes | c__Clostridia | o__Clostridiales | f__Lachnospiraceae | g__Butyrivibrio | s__Butyrivibrio_proteoclasticus | 22 |
| k__Bacteria | p__Firmicutes | c__Clostridia | o__Clostridiales | f__Lachnospiraceae | g__Catonella | s__Catonella_morbi | 22 |
| k__Bacteria | p__Firmicutes | c__Clostridia | o__Clostridiales | f__Lachnospiraceae | g__Coprococcus | s__Coprococcus_eutactus | 22 |
| k__Bacteria | p__Firmicutes | c__Clostridia | o__Clostridiales | f__Lachnospiraceae | g__Dorea | s__Dorea_sp._5-2 | 22 |
| k__Bacteria | p__Firmicutes | c__Clostridia | o__Clostridiales | f__Lachnospiraceae | g__Lachnobacterium | s__Lachnobacterium_bovis | 22 |
| k__Bacteria | p__Firmicutes | c__Clostridia | o__Clostridiales | f__Lachnospiraceae | g__Lachnoclostridium | s__Lachnoclostridium_phytofermentans | 22 |
| k__Bacteria | p__Firmicutes | c__Clostridia | o__Clostridiales | f__Lachnospiraceae | g__Lachnospira | s__Lachnospira_multipara | 22 |
| k__Bacteria | p__Firmicutes | c__Clostridia | o__Clostridiales | f__Lachnospiraceae | g__Oribacterium | s__Oribacterium_sp._P6A1 | 22 |
| k__Bacteria | p__Firmicutes | c__Clostridia | o__Clostridiales | f__Lachnospiraceae | g__Oribacterium | s__Oribacterium_sp._oral_taxon_078 | 22 |
| k__Bacteria | p__Firmicutes | c__Clostridia | o__Clostridiales | f__Lachnospiraceae | g__Pseudobutyrivibrio | s__Pseudobutyrivibrio_ruminis | 22 |
| k__Bacteria | p__Firmicutes | c__Clostridia | o__Clostridiales | f__Lachnospiraceae | g__Roseburia | s__Roseburia_faecis | 22 |
| k__Bacteria | p__Firmicutes | c__Clostridia | o__Clostridiales | f__Lachnospiraceae | g__Roseburia | s__Roseburia_intestinalis | 22 |
| k__Bacteria | p__Firmicutes | c__Clostridia | o__Clostridiales | f__Lachnospiraceae | g__Roseburia | s__Roseburia_inulinivorans | 22 |
| k__Bacteria | p__Firmicutes | c__Clostridia | o__Clostridiales | f__Lachnospiraceae | g__Shuttleworthia | s__Shuttleworthia_satelles | 22 |
| k__Bacteria | p__Firmicutes | c__Clostridia | o__Clostridiales | f__Lachnospiraceae | g__Lachnoanaerobaculum | s__Lachnoanaerobaculum_saburreum | 23 |
| k__Bacteria | p__Firmicutes | c__Clostridia | o__Clostridiales | f__Lachnospiraceae | g__Lachnoclostridium | s__[Clostridium]_saccharolyticum | 23 |
| k__Bacteria | p__Firmicutes | c__Clostridia | o__Clostridiales | f__Lachnospiraceae | g__Stomatobaculum | s__Stomatobaculum_longum | 23 |
| k__Bacteria | p__Firmicutes | c__Clostridia | o__Clostridiales | f__Peptostreptococca | g__Peptostreptococcus | s__Peptostreptococcus_anaerobius | 23 |
| k__Bacteria | p__Firmicutes | c__Clostridia | o__Clostridiales | f__Ruminococcaceae | g__Faecalibacterium | s__Faecalibacterium_prausnitzii | 23 |
| k__Bacteria | p__Firmicutes | c__Clostridia | o__Clostridiales | f__Ruminococcaceae | g__Ruminococcus | s__Ruminococcus_albus | 24 |
| k__Bacteria | p__Firmicutes | c__Clostridia | o__Clostridiales | f__Ruminococcaceae | g__Ruminococcus | s__Ruminococcus_bicirculans | 24 |
| k__Bacteria | p__Firmicutes | c__Clostridia | o__Clostridiales | f__Ruminococcaceae | g__Ruminococcus | s__Ruminococcus_callidus | 24 |
| k__Bacteria | p__Firmicutes | c__Clostridia | o__Clostridiales | f__Ruminococcaceae | g__Ruminococcus | s__Ruminococcus_flavefaciens | 24 |
| k__Bacteria | p__Firmicutes | c__Clostridia | o__Clostridiales | f__Ruminococcaceae | g__Ruminococcus | s__Ruminococcus_sp._FC2018 | 24 |
| k__Bacteria | p__Firmicutes | c__Clostridia | o__Clostridiales | f__Ruminococcaceae | g__Ruminococcus | s__Ruminococcus_sp._HUN007 | 24 |
| k__Bacteria | p__Firmicutes | c__Clostridia | o__Clostridiales | f__Ruminococcaceae | g__Ruminococcus | s__Ruminococcus_sp._NK3A76 | 24 |
| k__Bacteria | p__Firmicutes | c__Negativicutes | o__Acidaminococcales | f__Acidaminococcace | g__Phascolarctobacterium | s__Phascolarctobacterium_succinatutens | 25 |
| k__Bacteria | p__Firmicutes | c__Negativicutes | o__Veillonellales | f__Veillonellaceae | g__Dialister | s__Dialister_invisus | 25 |

**Supplementary Table 6. The regression coefficient of Baseline models**

| Model (Baseline) | Variables | Coefficient | Domain | Phylum | Class | Order | Family | Genus |
|---|---|---|---|---|---|---|---|---|
| Baseline Age + Sex | Age | 0,1200 | | | | | | |
| Baseline Age + Sex | Sex | 0,2471 | | | | | | |
| Baseline Covariates | Age | 0,1080 | | | | | | |
| Baseline Covariates | Body Mass Index | 0,0862 | | | | | | |
| Baseline Covariates | Smoking | 0,4921 | | | | | | |
| Baseline Covariates | Blood Pressure Treatment | 0,3742 | | | | | | |
| Baseline Covariates | Prevalent Diabetes | 0,8151 | | | | | | |
| Baseline Covariates | Prevalent CHD | 0,6245 | | | | | | |
| Baseline Covariates | Systolic Blood Pressure | 0,0087 | | | | | | |
| Baseline Covariates | NonHDL Cholesterol | -0,0830 | | | | | | |
| Baseline Covariates | Sex | 0,2871 | | | | | | |
| Baseline All | Age | 0,1116 | | | | | | |
| Baseline All | Body Mass Index | 0,0976 | | | | | | |
| Baseline All | Blood Pressure Treatment | 0,4139 | | | | | | |
| Baseline All | NonHDL Cholesterol | -0,0359 | | | | | | |
| Baseline All | Prevalent CHD | 0,5948 | | | | | | |
| Baseline All | Prevalent Diabetes | 0,6635 | | | | | | |
| Baseline All | Sex | 0,2635 | | | | | | |
| Baseline All | Smoking | 0,3377 | | | | | | |
| Baseline All | Systolic Blood Pressure | 0,0098 | | | | | | |
| Baseline All | *Methanobrevibacter smithii* | 0,0005 | Archaea | Euryarchaeota | Methanobacteria | Methanobacteriales | Methanobacteriaceae | Methanobrevibacter |
| Baseline All | *Eggerthella lenta* | 0,0324 | Bacteria | Actinobacteria | Coriobacteriia | Eggerthellales | Eggerthellaceae | Eggerthella |
| Baseline All | *Ruminococcus lactaris* | -0,1290 | Bacteria | Firmicutes | Clostridia | Clostridiales | Ruminococcaceae | Ruminococcus |
| Baseline All | *Ruthenibacterium lactatiform* | 0,0409 | Bacteria | Firmicutes | Clostridia | Clostridiales | Ruminococcaceae | Ruthenibacterium |
| Baseline All | *Subdoligranulum variabile* | -0,1020 | Bacteria | Firmicutes | Clostridia | Clostridiales | Ruminococcaceae | Subdoligranulum |
| Baseline All | *Catenibacterium mitsuokai* | 0,0933 | Bacteria | Firmicutes | Erysipelotrichia | Erysipelotrichales | Erysipelotrichaceae | Catenibacterium |
| Baseline All | *Coprobacillus sp. D6* | -0,1587 | Bacteria | Firmicutes | Erysipelotrichia | Erysipelotrichales | Erysipelotrichaceae | Coprobacillus |
| Baseline All | *[Clostridium] innocuum* | 0,0537 | Bacteria | Firmicutes | Erysipelotrichia | Erysipelotrichales | Erysipelotrichaceae | Erysipelatoclostridium |
| Baseline All | *Faecalitalea cylindroides* | 0,0072 | Bacteria | Firmicutes | Erysipelotrichia | Erysipelotrichales | Erysipelotrichaceae | Faecalitalea |
| Baseline All | *Holdemanella biformis* | 0,0288 | Bacteria | Firmicutes | Erysipelotrichia | Erysipelotrichales | Erysipelotrichaceae | Holdemanella |
| Baseline All | *Phascolarctobacterium succin* | 0,0684 | Bacteria | Firmicutes | Negativicutes | Acidaminococcales | Acidaminococcaceae | Phascolarctobacterium |
| Baseline All | *Dialister invisus* | 0,0271 | Bacteria | Firmicutes | Negativicutes | Veillonellales | Veillonellaceae | Dialister |
| Baseline All | *Bacteroides barnesiae* | -0,1328 | Bacteria | Bacteroidetes | Bacteroidia | Bacteroidales | Bacteroidaceae | Bacteroides |
| Baseline All | *Parasutterella excrementihomi* | -0,0584 | Bacteria | Proteobacteria | Betaproteobacteria | Burkholderiales | Sutterellaceae | Parasutterella |
| Baseline All | *Sutterella wadsworthensis* | -0,0470 | Bacteria | Proteobacteria | Betaproteobacteria | Burkholderiales | Sutterellaceae | Sutterella |
| Baseline All | *Bilophila wadsworthia* | -0,0275 | Bacteria | Proteobacteria | Deltaproteobacteria | Desulfovibrionales | Desulfovibrionaceae | Bilophila |
| Baseline All | *Desulfovibrio piger* | -0,0391 | Bacteria | Proteobacteria | Deltaproteobacteria | Desulfovibrionales | Desulfovibrionaceae | Desulfovibrio |
| Baseline All | *Escherichia coli* | -0,0744 | Bacteria | Proteobacteria | Gammaproteobacteria | Enterobacterales | Enterobacteriaceae | Escherichia |
| Baseline All | *Shigella dysenteriae* | 0,0561 | Bacteria | Proteobacteria | Gammaproteobacteria | Enterobacterales | Enterobacteriaceae | Shigella |
| Baseline All | *Shigella flexneri* | 0,0219 | Bacteria | Proteobacteria | Gammaproteobacteria | Enterobacterales | Enterobacteriaceae | Shigella |
| Baseline All | *Akkermansia muciniphila* | -0,0349 | Bacteria | Verrucomicrobia | Verrucomicrobiae | Verrucomicrobiales | Akkermansiaceae | Akkermansia |
| Baseline All | *Bacteroides thetaiotaomicron* | 0,0517 | BacteriaPlasmid | Bacteroidetes | Bacteroidia | Bacteroidales | Bacteroidaceae | Bacteroides |
| Baseline All | *[Eubacterium] eligens* | -0,0979 | BacteriaPlasmid | Firmicutes | Clostridia | Clostridiales | Eubacteriaceae | Eubacterium |
| Baseline All | *Bacteroides cellulosilyticus* | -0,0436 | Bacteria | Bacteroidetes | Bacteroidia | Bacteroidales | Bacteroidaceae | Bacteroides |
| Baseline All | *dsDNA viruses* | -0,0187 | Viruses | dsDNA viruses | Caudovirales | | | |
| Baseline All | *Bacteroides coprocola* | 0,0039 | Bacteria | Bacteroidetes | Bacteroidia | Bacteroidales | Bacteroidaceae | Bacteroides |
| Baseline All | *Bacteroides coprophilus* | 0,0119 | Bacteria | Bacteroidetes | Bacteroidia | Bacteroidales | Bacteroidaceae | Bacteroides |
| Baseline All | *Bacteroides faecichinchillae* | -0,0796 | Bacteria | Bacteroidetes | Bacteroidia | Bacteroidales | Bacteroidaceae | Bacteroides |
| Baseline All | *Bacteroides fluxus* | -0,0026 | Bacteria | Bacteroidetes | Bacteroidia | Bacteroidales | Bacteroidaceae | Bacteroides |
| Baseline All | *Bacteroides fragilis* | 0,0796 | Bacteria | Bacteroidetes | Bacteroidia | Bacteroidales | Bacteroidaceae | Bacteroides |
| Baseline All | *Bacteroides oleiciplenus* | -0,1462 | Bacteria | Bacteroidetes | Bacteroidia | Bacteroidales | Bacteroidaceae | Bacteroides |
| Baseline All | *Bacteroides ovatus* | 0,0028 | Bacteria | Bacteroidetes | Bacteroidia | Bacteroidales | Bacteroidaceae | Bacteroides |
| Baseline All | *Bifidobacterium adolescentis* | -0,0297 | Bacteria | Actinobacteria | Actinobacteria | Bifidobacteriales | Bifidobacteriaceae | Bifidobacterium |
| Baseline All | *Bacteroides plebeius* | 0,0103 | Bacteria | Bacteroidetes | Bacteroidia | Bacteroidales | Bacteroidaceae | Bacteroides |
| Baseline All | *Bacteroides salanitronis* | 0,0185 | Bacteria | Bacteroidetes | Bacteroidia | Bacteroidales | Bacteroidaceae | Bacteroides |
| Baseline All | *Bacteroides salyersiae* | 0,0538 | Bacteria | Bacteroidetes | Bacteroidia | Bacteroidales | Bacteroidaceae | Bacteroides |
| Baseline All | *Bacteroides stercoris* | -0,1386 | Bacteria | Bacteroidetes | Bacteroidia | Bacteroidales | Bacteroidaceae | Bacteroides |
| Baseline All | *Bacteroides thetaiotaomicron* | 0,0273 | Bacteria | Bacteroidetes | Bacteroidia | Bacteroidales | Bacteroidaceae | Bacteroides |
| Baseline All | *Bacteroides uniformis* | -0,0588 | Bacteria | Bacteroidetes | Bacteroidia | Bacteroidales | Bacteroidaceae | Bacteroides |
| Baseline All | *Bacteroides vulgatus* | 0,0130 | Bacteria | Bacteroidetes | Bacteroidia | Bacteroidales | Bacteroidaceae | Bacteroides |
| Baseline All | *Barnesiella intestinihominis* | 0,0680 | Bacteria | Bacteroidetes | Bacteroidia | Bacteroidales | Barnesiellaceae | Barnesiella |
| Baseline All | *Coprobacter fastidiosus* | -0,0004 | Bacteria | Bacteroidetes | Bacteroidia | Bacteroidales | Barnesiellaceae | Coprobacter |
| Baseline All | *Coprobacter secundus* | -0,0138 | Bacteria | Bacteroidetes | Bacteroidia | Bacteroidales | Barnesiellaceae | Coprobacter |
| Baseline All | *Bifidobacterium bifidum* | 0,0167 | Bacteria | Actinobacteria | Actinobacteria | Bifidobacteriales | Bifidobacteriaceae | Bifidobacterium |
| Baseline All | *Butyricimonas virosa* | -0,0469 | Bacteria | Bacteroidetes | Bacteroidia | Bacteroidales | Odoribacteraceae | Butyricimonas |
| Baseline All | *Odoribacter splanchnicus* | 0,0212 | Bacteria | Bacteroidetes | Bacteroidia | Bacteroidales | Odoribacteraceae | Odoribacter |
| Baseline All | *Paraprevotella xylaniphila* | 0,0241 | Bacteria | Bacteroidetes | Bacteroidia | Bacteroidales | Prevotellaceae | Paraprevotella |
| Baseline All | *Prevotella bivia* | -0,0949 | Bacteria | Bacteroidetes | Bacteroidia | Bacteroidales | Prevotellaceae | Prevotella |
| Baseline All | *Prevotella copri* | 0,0346 | Bacteria | Bacteroidetes | Bacteroidia | Bacteroidales | Prevotellaceae | Prevotella |
| Baseline All | *Prevotella sp. 109* | 0,0604 | Bacteria | Bacteroidetes | Bacteroidia | Bacteroidales | Prevotellaceae | Prevotella |
| Baseline All | *Prevotella sp. P4-76* | 0,0641 | Bacteria | Bacteroidetes | Bacteroidia | Bacteroidales | Prevotellaceae | Prevotella |
| Baseline All | *Prevotella stercorea* | 0,0110 | Bacteria | Bacteroidetes | Bacteroidia | Bacteroidales | Prevotellaceae | Prevotella |
| Baseline All | *Prevotella timonensis* | 0,0778 | Bacteria | Bacteroidetes | Bacteroidia | Bacteroidales | Prevotellaceae | Prevotella |
| Baseline All | *Alistipes finegoldii* | 0,0168 | Bacteria | Bacteroidetes | Bacteroidia | Bacteroidales | Rikenellaceae | Alistipes |
| Baseline All | *Bifidobacterium breve* | 0,1081 | Bacteria | Actinobacteria | Actinobacteria | Bifidobacteriales | Bifidobacteriaceae | Bifidobacterium |
| Baseline All | *Alistipes ihumii* | 0,0249 | Bacteria | Bacteroidetes | Bacteroidia | Bacteroidales | Rikenellaceae | Alistipes |
| Baseline All | *Alistipes indistinctus* | -0,0494 | Bacteria | Bacteroidetes | Bacteroidia | Bacteroidales | Rikenellaceae | Alistipes |
| Baseline All | *Alistipes inops* | -0,0165 | Bacteria | Bacteroidetes | Bacteroidia | Bacteroidales | Rikenellaceae | Alistipes |
| Baseline All | *Alistipes obesi* | 0,0197 | Bacteria | Bacteroidetes | Bacteroidia | Bacteroidales | Rikenellaceae | Alistipes |
| Baseline All | *Alistipes putredinis* | 0,0651 | Bacteria | Bacteroidetes | Bacteroidia | Bacteroidales | Rikenellaceae | Alistipes |
| Baseline All | *Alistipes senegalensis* | 0,0368 | Bacteria | Bacteroidetes | Bacteroidia | Bacteroidales | Rikenellaceae | Alistipes |
| Baseline All | *Alistipes shahii* | -0,1131 | Bacteria | Bacteroidetes | Bacteroidia | Bacteroidales | Rikenellaceae | Alistipes |
| Baseline All | *Alistipes timonensis* | 0,0499 | Bacteria | Bacteroidetes | Bacteroidia | Bacteroidales | Rikenellaceae | Alistipes |
| Baseline All | *Parabacteroides distasonis* | 0,0385 | Bacteria | Bacteroidetes | Bacteroidia | Bacteroidales | Tannerellaceae | Parabacteroides |
| Baseline All | *Parabacteroides goldsteinii* | 0,0382 | Bacteria | Bacteroidetes | Bacteroidia | Bacteroidales | Tannerellaceae | Parabacteroides |
| Baseline All | *Bifidobacterium longum* | -0,1198 | Bacteria | Actinobacteria | Actinobacteria | Bifidobacteriales | Bifidobacteriaceae | Bifidobacterium |
| Baseline All | *Parabacteroides johnsonii* | 0,1330 | Bacteria | Bacteroidetes | Bacteroidia | Bacteroidales | Tannerellaceae | Parabacteroides |
| Baseline All | *Streptococcus salivarius* | -0,0649 | Bacteria | Firmicutes | Bacilli | Lactobacillales | Streptococcaceae | Streptococcus |
| Baseline All | *Streptococcus thermophilus* | 0,0222 | Bacteria | Firmicutes | Bacilli | Lactobacillales | Streptococcaceae | Streptococcus |
| Baseline All | *Butyricicoccus pullicaecorum* | -0,2007 | Bacteria | Firmicutes | Clostridia | Clostridiales | Clostridiaceae | Butyricicoccus |
| Baseline All | *Clostridium phoceensis* | -0,1297 | Bacteria | Firmicutes | Clostridia | Clostridiales | Clostridiaceae | Clostridium |
| Baseline All | *Clostridium sp. L2-50* | -0,1116 | Bacteria | Firmicutes | Clostridia | Clostridiales | Clostridiaceae | Clostridium |
| Baseline All | *Clostridium sp. M62/1* | 0,2568 | Bacteria | Firmicutes | Clostridia | Clostridiales | Clostridiaceae | Clostridium |
| Baseline All | *Hungatella hathewayi* | 0,0283 | Bacteria | Firmicutes | Clostridia | Clostridiales | Clostridiaceae | Hungatella |
| Baseline All | *Eubacterium ramulus* | -0,0021 | Bacteria | Firmicutes | Clostridia | Clostridiales | Eubacteriaceae | Eubacterium |
| Baseline All | *Eubacterium ventriosum* | -0,0730 | Bacteria | Firmicutes | Clostridia | Clostridiales | Eubacteriaceae | Eubacterium |
| Baseline All | *Cellulomonas carbonis* | -0,0491 | Bacteria | Actinobacteria | Actinobacteria | Micrococcales | Cellulomonadaceae | Cellulomonas |
| Baseline All | *[Eubacterium] eligens* | 0,0811 | Bacteria | Firmicutes | Clostridia | Clostridiales | Eubacteriaceae | Eubacterium |
| Baseline All | *[Eubacterium] hallii* | 0,0667 | Bacteria | Firmicutes | Clostridia | Clostridiales | Eubacteriaceae | Eubacterium |
| Baseline All | *Anaerostipes hadrus* | 0,0999 | Bacteria | Firmicutes | Clostridia | Clostridiales | Lachnospiraceae | Anaerostipes |
| Baseline All | *Blautia hansenii* | 0,0666 | Bacteria | Firmicutes | Clostridia | Clostridiales | Lachnospiraceae | Blautia |
| Baseline All | *Blautia obeum* | -0,1597 | Bacteria | Firmicutes | Clostridia | Clostridiales | Lachnospiraceae | Blautia |
| Baseline All | *Blautia sp. KLE 1732* | 0,0005 | Bacteria | Firmicutes | Clostridia | Clostridiales | Lachnospiraceae | Blautia |
| Baseline All | *[Ruminococcus] gnavus* | 0,0248 | Bacteria | Firmicutes | Clostridia | Clostridiales | Lachnospiraceae | Blautia |
| Baseline All | *[Ruminococcus] torques* | -0,0337 | Bacteria | Firmicutes | Clostridia | Clostridiales | Lachnospiraceae | Blautia |
| Baseline All | *Butyrivibrio crossotus* | 0,0262 | Bacteria | Firmicutes | Clostridia | Clostridiales | Lachnospiraceae | Butyrivibrio |
| Baseline All | *Coprococcus comes* | -0,0597 | Bacteria | Firmicutes | Clostridia | Clostridiales | Lachnospiraceae | Coprococcus |
| Baseline All | *Collinsella aerofaciens* | 0,0370 | Bacteria | Actinobacteria | Coriobacteriia | Coriobacteriales | Coriobacteriaceae | Collinsella |
| Baseline All | *Coprococcus eutactus* | -0,1180 | Bacteria | Firmicutes | Clostridia | Clostridiales | Lachnospiraceae | Coprococcus |
| Baseline All | *Dorea formicigenerans* | 0,1063 | Bacteria | Firmicutes | Clostridia | Clostridiales | Lachnospiraceae | Dorea |
| Baseline All | *Dorea longicatena* | -0,2837 | Bacteria | Firmicutes | Clostridia | Clostridiales | Lachnospiraceae | Dorea |
| Baseline All | *Dorea sp. 5-2* | -0,0318 | Bacteria | Firmicutes | Clostridia | Clostridiales | Lachnospiraceae | Dorea |
| Baseline All | *Faecalicatena contorta* | 0,0382 | Bacteria | Firmicutes | Clostridia | Clostridiales | Lachnospiraceae | Faecalicatena |
| Baseline All | *[Clostridium] asparagiforme* | -0,0979 | Bacteria | Firmicutes | Clostridia | Clostridiales | Lachnospiraceae | Lachnoclostridium |
| Baseline All | *[Clostridium] bolteae* | -0,0259 | Bacteria | Firmicutes | Clostridia | Clostridiales | Lachnospiraceae | Lachnoclostridium |
| Baseline All | *[Clostridium] citroniae* | -0,0532 | Bacteria | Firmicutes | Clostridia | Clostridiales | Lachnospiraceae | Lachnoclostridium |
| Baseline All | *[Clostridium] glycyrrhizinilytic* | 0,3094 | Bacteria | Firmicutes | Clostridia | Clostridiales | Lachnospiraceae | Lachnoclostridium |
| Baseline All | *[Clostridium] symbiosum* | 0,1960 | Bacteria | Firmicutes | Clostridia | Clostridiales | Lachnospiraceae | Lachnoclostridium |
| Baseline All | *Senegalimassilia anaerobia* | 0,0800 | Bacteria | Actinobacteria | Coriobacteriia | Coriobacteriales | Coriobacteriaceae | Senegalimassilia |
| Baseline All | *Roseburia faecis* | -0,0411 | Bacteria | Firmicutes | Clostridia | Clostridiales | Lachnospiraceae | Roseburia |
| Baseline All | *Roseburia hominis* | -0,0622 | Bacteria | Firmicutes | Clostridia | Clostridiales | Lachnospiraceae | Roseburia |
| Baseline All | *Roseburia intestinalis* | -0,0096 | Bacteria | Firmicutes | Clostridia | Clostridiales | Lachnospiraceae | Roseburia |
| Baseline All | *Roseburia inulinivorans* | 0,1615 | Bacteria | Firmicutes | Clostridia | Clostridiales | Lachnospiraceae | Roseburia |
| Baseline All | *Sellimonas intestinalis* | -0,1135 | Bacteria | Firmicutes | Clostridia | Clostridiales | Lachnospiraceae | Sellimonas |
| Baseline All | *Tyzzerella nexilis* | 0,0151 | Bacteria | Firmicutes | Clostridia | Clostridiales | Lachnospiraceae | Tyzzerella |
| Baseline All | *[Clostridium] lactatifermentan* | 0,1447 | Bacteria | Firmicutes | Clostridia | Clostridiales | Lachnospiraceae | Tyzzerella |
| Baseline All | *Oscillibacter sp. 1-3* | -0,2322 | Bacteria | Firmicutes | Clostridia | Clostridiales | Oscillospiraceae | Oscillibacter |
| Baseline All | *Oscillibacter sp. ER4* | -0,0359 | Bacteria | Firmicutes | Clostridia | Clostridiales | Oscillospiraceae | Oscillibacter |
| Baseline All | *Oscillibacter sp. KLE 1745* | -0,0327 | Bacteria | Firmicutes | Clostridia | Clostridiales | Oscillospiraceae | Oscillibacter |
| Baseline All | *Adlercreutzia equolifaciens* | 0,0690 | Bacteria | Actinobacteria | Coriobacteriia | Eggerthellales | Eggerthellaceae | Adlercreutzia |
| Baseline All | *Intestinibacter bartlettii* | -0,0372 | Bacteria | Firmicutes | Clostridia | Clostridiales | Peptostreptococcaceae | Intestinibacter |
| Baseline All | *Anaerotruncus colihominis* | 0,0935 | Bacteria | Firmicutes | Clostridia | Clostridiales | Ruminococcaceae | Anaerotruncus |
| Baseline All | *Faecalibacterium prausnitzii* | 0,3259 | Bacteria | Firmicutes | Clostridia | Clostridiales | Ruminococcaceae | Faecalibacterium |
| Baseline All | *Fournierella massiliensis* | 0,0241 | Bacteria | Firmicutes | Clostridia | Clostridiales | Ruminococcaceae | Fournierella |
| Baseline All | *[Clostridium] leptum* | 0,0610 | Bacteria | Firmicutes | Clostridia | Clostridiales | Ruminococcaceae | Ruminiclostridium |
| Baseline All | *[Eubacterium] siraeum* | 0,0999 | Bacteria | Firmicutes | Clostridia | Clostridiales | Ruminococcaceae | Ruminiclostridium |
| Baseline All | *Ruminococcus bicirculans* | 0,1013 | Bacteria | Firmicutes | Clostridia | Clostridiales | Ruminococcaceae | Ruminococcus |
| Baseline All | *Ruminococcus callidus* | 0,0189 | Bacteria | Firmicutes | Clostridia | Clostridiales | Ruminococcaceae | Ruminococcus |
| Baseline All | *Ruminococcus champanellensi* | -0,0458 | Bacteria | Firmicutes | Clostridia | Clostridiales | Ruminococcaceae | Ruminococcus |
| Baseline All | *Ruminococcus faecis* | -0,2803 | Bacteria | Firmicutes | Clostridia | Clostridiales | Ruminococcaceae | Ruminococcus |

**Supplementary Table 7. Post challenge model evaluation**

| Model combinations | Harrel's C index | Hosmer Lemeshow P-value |
|---|---|---|
| **SB2** | 0,83440000 | 0,0018 |
| **DenverFINRISKHacky** | 0,82710000 | 0,0092 |
| **SB2 DFH** | 0,83690000 | 0,1551 |
| **SB2 DFH YG-HZ team** | 0,83990000 | 0,0699 |
| **SB2 DFH Metformin-121** | 0,84010000 | 2,13E-06 |
| **SB2 DFH TristanF** | 0,83670000 | 2,13E-06 |
| **SB2 DFH YG-HZ team Metformin-121** | 0,84010000 | 2,73E-32 |
| **SB2 DFH YG-HZ team Metformin-121 TristanF** | 0,84110000 | 1,94E-34 |
| **SB2 DFH YG-HZ team Metformin-121 TristanF UTKteam** | 0,83900000 | 7,12E-40 |
| **SB2 DFH YG-HZ team Metformin-121 TristanF UTKteam Pteam** | 0,83770000 | 5,77E-98 |
| **SB2 final submission (without phylum)** | 0,83880000 | 0,0102 |
| **SB2 Age unpenalized (without phylum)** | 0,8392 | 0,0019 |
| **SB2-HF strict definition** | 0,8541 | 0,1358 |
| **DFH-HF strict definition** | 0,8454 | 4,53E-11 |

**Supplementary Table 8. Comparison of bootstrapped evaluation metric (n=1000) relative to the top performer**

| Team A | Team B | Harrell's C Bayes Factor | Hosmer-Lemeshow Bayes Factor |
|---|---|---|---|
| SB2 | Baseline All Covariat | 1,67 | 7,13 |
| SB2 | DFH | 6,09 | 1,46 |
| SB2 | TristanF | 2,53 | >500 |
| SB2 | YG-HZ team | 5,1 | >500 |
| SB2 | Metformin-121 | 33,8 | >500 |
| SB2 | Baseline All | 57,82 | 15,95 |
| SB2 | Baseline Age-Sex | 332,33 | 165,67 |
| SB2 | UTKteam | >500 | >500 |
| SB2 | Pteam | >500 | >500 |
